# Trends in the conduct and reporting of clinical prediction model development and validation: a systematic review

**DOI:** 10.1101/2021.10.22.21265374

**Authors:** Cynthia Yang, Jan A. Kors, Solomon Ioannou, Luis H. John, Aniek F. Markus, Alexandros Rekkas, Maria A.J. de Ridder, Tom M. Seinen, Ross D. Williams, Peter R. Rijnbeek

## Abstract

**Objectives:** This systematic review aims to provide further insights into the conduct and reporting of clinical prediction model development and validation over time. We focus on assessing the reporting of information necessary to enable external validation by other investigators.

**Materials and Methods:** We searched Embase, Medline, Web-of-Science, Cochrane Library and Google Scholar to identify studies that developed one or more multivariable prognostic prediction models using electronic health record (EHR) data published in the period 2009-2019.

**Results:** We identified 422 studies that developed a total of 579 clinical prediction models using EHR data. We observed a steep increase over the years in the number of developed models. The percentage of models externally validated in the same paper remained at around 10%. Throughout 2009-2019, for both the target population and the outcome definitions, code lists were provided for less than 20% of the models. For about half of the models that were developed using regression analysis, the final model was not completely presented.

**Discussion:** Overall, we observed limited improvement over time in the conduct and reporting of clinical prediction model development and validation. In particular, the prediction problem definition was often not clearly reported, and the final model was often not completely presented.

**Conclusion:** Improvement in the reporting of information necessary to enable external validation by other investigators is still urgently needed to increase clinical adoption of developed models.

## INTRODUCTION

The wide implementation of the electronic health record (EHR) in recent decades drastically increased the availability of patient-level data for clinical prediction modelling. This has led to the development of many clinical prediction models using EHR data. Still, very few developed models are adopted in clinical practice. A reason for this lack of adoption is that before implementing a prediction model in clinical practice, it is important to ensure generalizability and robustness of the model’s prediction performance; this can be achieved by externally validating the model across various databases.[1-3] However, very few developed models have been externally validated by the original investigators.[1, 4, 5] As a result, for most developed models it remains unclear whether the prediction performance is generalizable and robust. It should also be possible for other investigators to perform external validation of a developed model.[3] A prerequisite for this is good conduct and reporting of model development and validation in the original study. In particular, the prediction problem definition needs to be clearly reported and the final model needs to be completely presented.[6] A prediction problem definition consists of several components that are specified as follows in our previously published standardized prediction framework:[7] among a *target population* of patients at an index date, predict which patients will experience some *outcome* during a *time-at-risk* period. Prediction is then done using predictors that are constructed using information from an *observation window* prior to the index date.

Previous systematic reviews on the development and validation of clinical prediction models covered different periods prior to 2015.[1, 4, 5] They all highlighted a number of improvements to allow for better interpretation of the presented results by other investigators: how missing data were handled should always explicitly be mentioned, model calibration should be assessed, and external validation should be performed. To encourage improvement in the conduct and reporting of model development and validation, the Transparent Reporting of a multivariable clinical prediction model for Individual Prognosis or Diagnosis (TRIPOD) Statement was published in January 2015.[8] A recent study assessed the adherence to the TRIPOD Statement in clinical prediction studies published in a selection of high-impact general medicine journals.[9] Their results suggested no significant impact of TRIPOD on the overall reporting in the two years following introduction. However, their study only covered studies up to 2017 and their selection of studies might not be representative of the entire field. No previous systematic review has assessed the trends in the conduct and reporting of model development and validation in the field of clinical prediction modelling over a longer period. Additionally, no previous systematic review has specifically assessed the reporting of information necessary to enable external validation by other investigators.

This systematic review aims to provide further insights into the conduct and reporting of clinical prediction model development and validation over time. We focus on assessing the reporting of information necessary to enable external validation by other investigators in studies that developed models using EHR data published in the period 2009-2019.

## METHODS

For reporting our systematic review, we followed the Preferred Reporting Items for Systematic Review and Meta-Analyses (PRISMA) guidelines.[10]

### Data sources and searches

To identify relevant papers, we searched Embase, Medline, Web-of-Science, Cochrane Library and Google Scholar. Information specialists with expert knowledge of medical terminology and databases were consulted to formulate the search queries (Appendix A). The search was limited to papers written in English and published in the period 2009-2019. Animal studies and studies that were not original research (e.g., comments, letters, editorials) or had no abstract were excluded. The search was performed on November 15, 2019.

### Study selection

We included all papers (including conference proceedings) that described the development of one or more multivariable prediction models using EHR data to estimate a patient’s probability of a particular clinical outcome occurring within a certain period in the future (i.e., prognostic prediction). Papers for which any of the following holds were excluded:

- there was a methodological focus (e.g., focusing on methodological improvements)
- the primary aim was evaluating predictor associations instead of model development
- only simulated data were used
- the study was a review of the literature
- we were unable to obtain the full text

One reviewer (C.Y.) screened all titles and abstracts to identify potentially eligible papers. The same reviewer then assessed eligibility of all remaining papers based on the full text.

### Data extraction and analysis

Data extraction was completed by multiple reviewers (J.A.K., S.I., L.H.J., A.F.M., A.R., M.A.J.R., T.M.S., R.D.W.) and verified by a second reviewer (C.Y.). Data extraction was based on the Checklist for critical Appraisal and data extraction for systematic Reviews of prediction Modelling Studies (CHARMS) Checklist,[11] and the TRIPOD Statement.[8] The data extraction form contained items from these two checklists together with some additional items (e.g., the reporting of code lists to define the clinical prediction problem).

Data were extracted from the abstract, main text, and any available supplemental material. We extracted data on several domains for each model in each study as follows.

‐ Data origin: the country from which the EHR data used originated.
‐ Data characteristics: the number of observations, the number of outcome events in the development dataset, and the number of candidate predictors.
‐ Data handling: the handling of missing data, the use of any class imbalance method.
‐ Modelling method: the type of algorithm used for model development.
‐ Prediction problem definition: whether inclusion/exclusion criteria for the target population were described, whether code lists to define the target population were provided, whether code lists to define the outcome were provided, whether the time-at-risk period was reported, whether all individual candidate predictors were listed, whether code lists to define the candidate predictors were provided, and whether the observation window for candidate predictor construction was reported.
‐ Final model presentation: the reported number of predictors in the final model, and whether the final model was completely presented. Depending on the modelling method, the final model can be completely presented using a full model equation (including intercept and coefficients), a simplified scoring system, a nomogram, an online tool, or a software package containing the analytical source code.[3]
‐ Model validation. We grouped each model into one of the following three categories: 1) externally validated, when performance was assessed on data from a database other than the development set, 2) internally validated only, when performance was assessed on the development set by split-sample, cross-validation, temporal validation, or bootstrapping, and 3) not validated, when performance was not assessed or only assessed on the same data that were used to train the model (referred to as the apparent performance). Prediction performance is typically characterized by evaluating a model’s calibration and discrimination.[12] Discrimination is usually assessed using a receiver operating characteristic (ROC) curve, with the area under the ROC curve (AUROC) as summary measure. Graphical assessment of calibration using a calibration plot is widely recommended.[13] From both internal and external validation results we therefore extracted the reported AUROC, whether the ROC curve was presented, whether a calibration plot was presented, and whether any other calibration measures (e.g., the calibration intercept and slope) were reported. In case of external validation, we additionally extracted: the number of observations, the number of outcome events, and whether data used for validation were from another country.

To investigate the trends in the period 2009-2019, we assessed the extracted data for the periods 2009-2014 and 2015-2019 separately.

## RESULTS

Our initial search resulted in a total of 9,942 papers. After duplicates were removed, 6,235 titles and abstracts were screened. From this, 1,075 potentially eligible papers were identified. Upon full-text inspection, 422 papers were eventually included for data extraction.

The study selection is presented in a PRISMA flow diagram (Appendix B). A reference list of the 422 included papers ordered by publication year is provided in Appendix C.

In total, we extracted data for 579 models from 422 studies (with 1-6 models per study). We observed a strong increase in the total number of models over the years (Table 1), with 135 models in 101 studies in the period 2009-2014 and 444 models in 321 studies in the period 2015-2019. The data used in these studies originated from EHRs in 38 different countries (13 countries in the period 2009-2014 and 35 countries in the period 2015-2019).

**Table 1.**
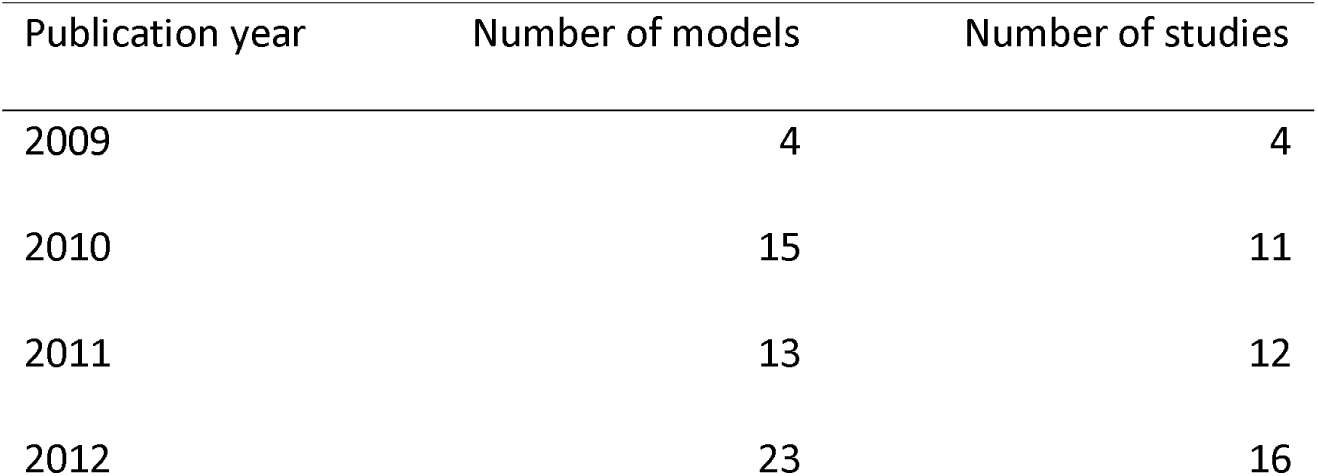
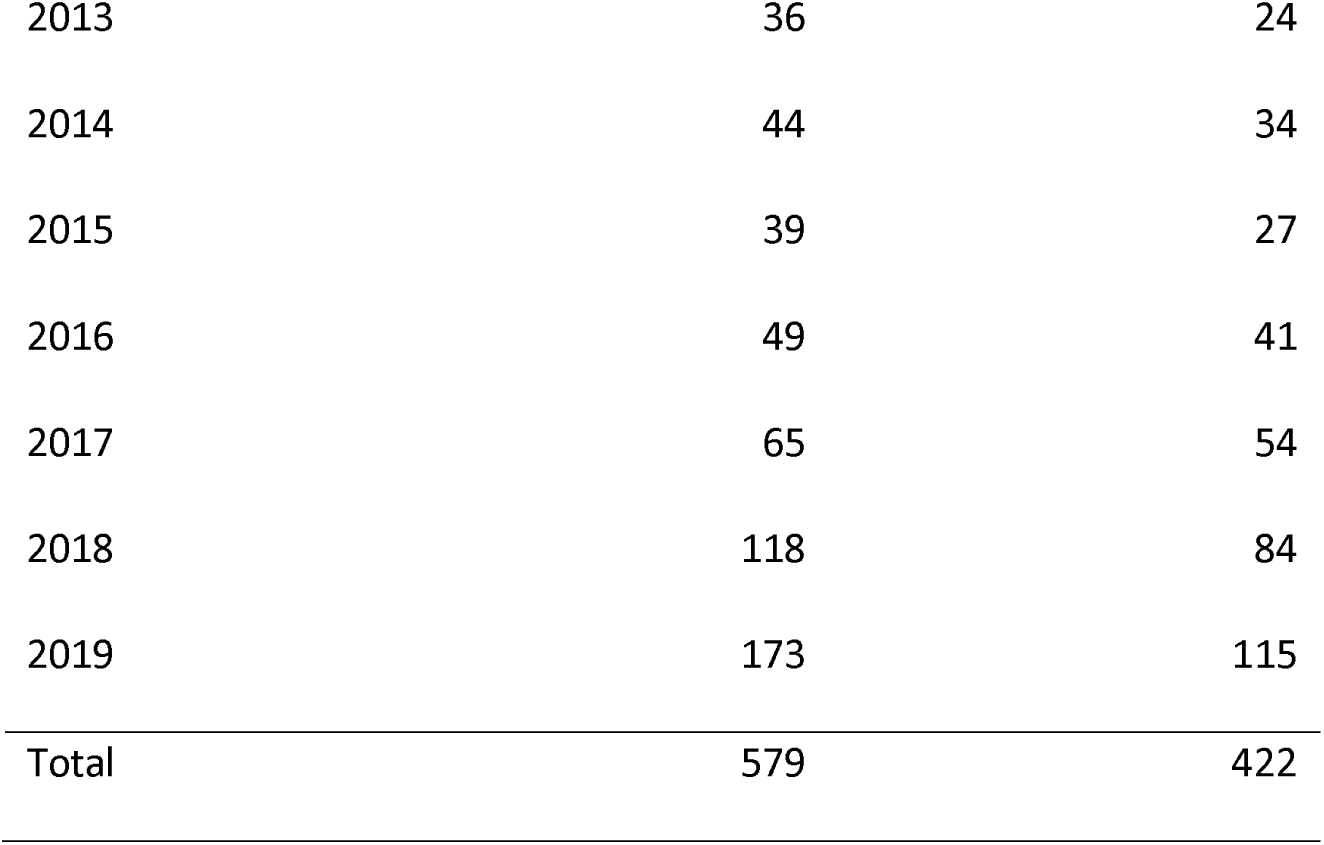
Trends in the publication of developed prediction models

| Publication year | Number of models | Number of studies |
| --- | --- | --- |
| 2009 | 4 | 4 |
| 2010 | 15 | 11 |
| 2011 | 13 | 12 |
| 2012 | 23 | 16 |
| 2013 | 36 | 24 |
| 2014 | 44 | 34 |
| 2015 | 39 | 27 |
| 2016 | 49 | 41 |
| 2017 | 65 | 54 |
| 2018 | 118 | 84 |
| 2019 | 173 | 115 |
| Total | 579 | 422 |

### Data characteristics

We investigated the characteristics of the EHR data used for model development. In both the periods 2009-2014 and 2015-2019, the number of observations in the development dataset was reported for 98% (132/135 and 433/444) of all models. The median reported number of observations increased from 7,086 (IQR: 1,293; 76,785) to 15,865 (IQR: 1,782; 68,319). The percentage of models for which the number of outcome events was explicitly reported slightly decreased from 90% (121/135) to 86% (383/444). The median reported number of outcome events increased from 536 (IQR: 199; 3,942) to 857 (IQR: 199; 4,135).

The percentage of models for which the number of candidate predictors was explicitly reported increased from 39% (53/135) to 50% (223/444). The median reported number of candidate predictors increased from 21 (IQR: 12; 300) to 46 (IQR: 23; 241).

### Data handling

The handling of missing data was reported for about half (61/135 and 224/444) of the models in both periods. An imputation method, an indicator method, or a combination of methods was applied for 72% (44/61 and 162/224) of these models. The reported use of class imbalance methods increased from 7% (9/135) to 13% (57/444) of the models.

### Modelling method

Various modelling methods were used for model development. We categorized these as Regression analysis, Decision tree learning, Ensemble method (e.g., when multiple decision trees are combined), Neural network (which includes deep learning), Bayesian network, Support vector machine, or Other (Figure 1). The percentage of models developed using regression analysis decreased from 76% (104/135) to 67% (300/444). Ensemble methods increased from 6% (8/135) to 19% (84/444) of the models, and neural networks increased from 1% (1/135) to 5% (24/444) of the models. Bayesian network decreased from 4% (5/135) to 2% (10/444) of the models. In both periods, 2% (3/135 and 9/444) of the models were developed using decision tree learning, and 1% (2/135 and 4/444) of the models were developed using a support vector machine. The Other category contained models for which the modelling method was unclear, models that were manually constructed, and a Hidden Markov model; this percentage decreased from 9% (12/135) to 3% (13/444).

**Figure 1.**
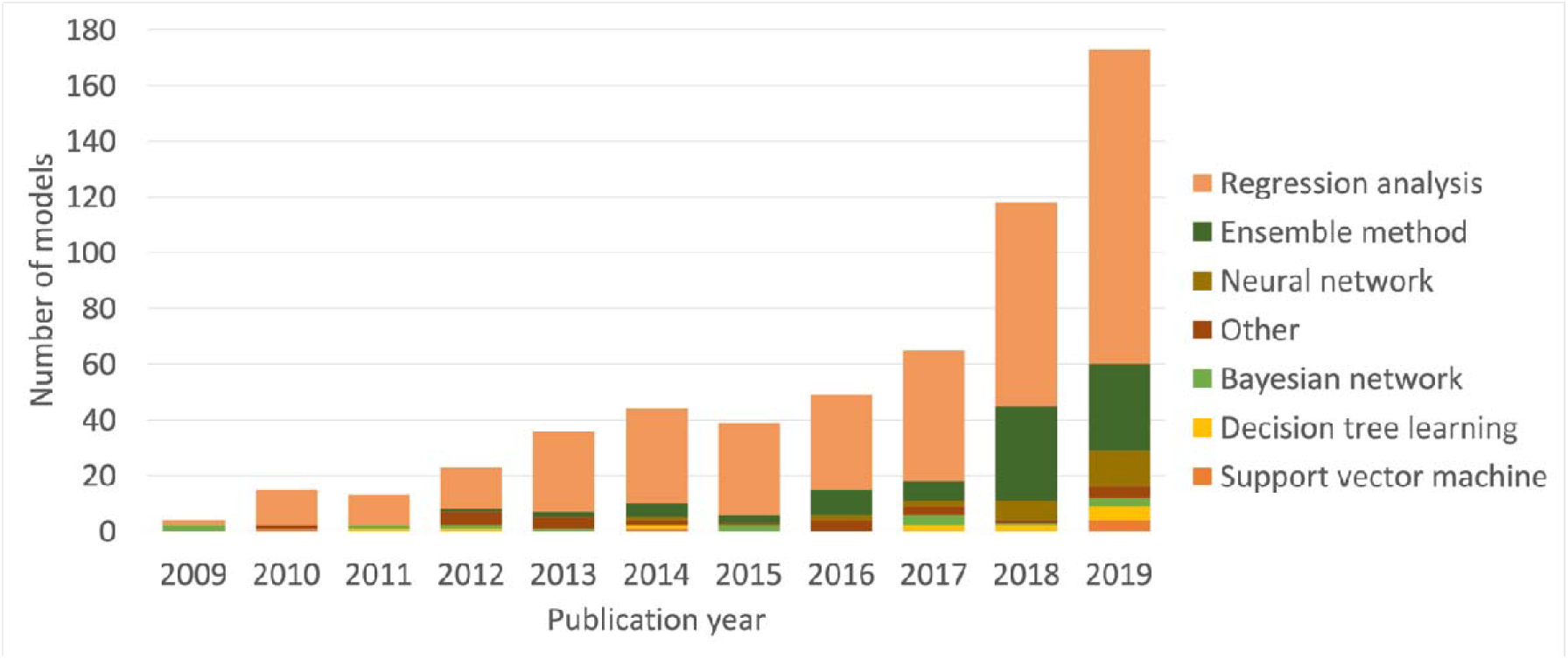
Trends in modelling methods

### Prediction problem definition

To evaluate the reporting of the prediction problem definition, we separately assessed each component that needs to be defined (Table 2). In both periods, for about 90% of the models the inclusion/exclusion criteria for the target population were described, while code lists to define the target population were provided for less than 20% of the models. Code lists to define the outcome were also provided for less than 20% of the models in both periods. The time-at-risk period was reported for 84% of the models in both periods.

**Table 2.** Trends in the reporting of definitions

| Component | 2009-2014 (N=135) | 2015-2019 (N=444) |
| --- | --- | --- |
| Target population – inclusion/exclusion criteria described (n, %) | 122 (90%) | 391 (88%) |
| Target population – provided through code list (n, %) | 19 (14%) | 81 (18%) |
| Outcome – provided through code list (n, %) | 22 (16%) | 81 (18%) |
| Time-at-risk – reported (n, %) | 114 (84%) | 375 (84%) |
| Candidate predictors – listed (n, %) | 91 (67%) | 301 (68%) |
| Candidate predictors – observation window reported (n, %) | 62 (46%) | 224 (50%) |
| Candidate predictors – provided through code list (n, %) | 13 (10%) | 46 (10%) |

In both periods, an overview of all candidate predictors was provided for two thirds of the models, while code lists to define the candidate predictors were provided for 10% of the models. The percentage of models for which the observation window for the construction of all candidate predictors was reported slightly increased from 46% to 50%.

### Final model presentation

We assessed the final model presentation for each modelling method category separately (Table 3). For Regression analysis, the final model was completely presented for about half of the models in both periods. For about two thirds (66/104 and 211/300) of the regression models in both periods, the number of predictors in the final model was explicitly reported. The median reported number of predictors in the final model slightly increased from 8 (IQR: 5; 15) to 10 (IQR: 6; 18). For more complex models (e.g., models with many predictors, ensemble models, or neural networks) it is often difficult or not possible to present a full model equation (including intercept and coefficients), a simplified scoring system, or a nomogram. Alternatively, one could resort to sharing an online tool or a software package containing the analytical source code. However, this was rare for the included studies.

**Table 3.** Trends in final model presentation

| Modelling method category | Final model completely presented in 2009-2014 (n, %) | Final model completely presented in 2015-2019 (n, %) |
| --- | --- | --- |
| Regression analysis (N=404) | 55 (53%) | 148 (49%) |
| Ensemble method (N=92) | 0 (0%) | 3 (4%) |
| Neural network (N=25) | 0 (0%) | 2 (8%) |
| Other (N=25) | 9 (75%) | 7 (54%) |
| Bayesian network (N=15) | 0 (0%) | 9 (40%) |
| Decision tree learning (N=12) | 1 (33%) | 7 (78%) |
| Support vector machine (N=6) | 0 (0%) | 0 (0%) |

### Model validation

External validation slightly increased from 10% (14/135) to 12% (55/444), internal validation only increased from 76% (103/135) to 81% (358/444), and no validation decreased from 13% (18/135) to 7% (31/444) (Figure 2). The percentage of externally validated models that were validated using data from another country remained less than 10% (1/14 and 5/55).

**Figure 2.**
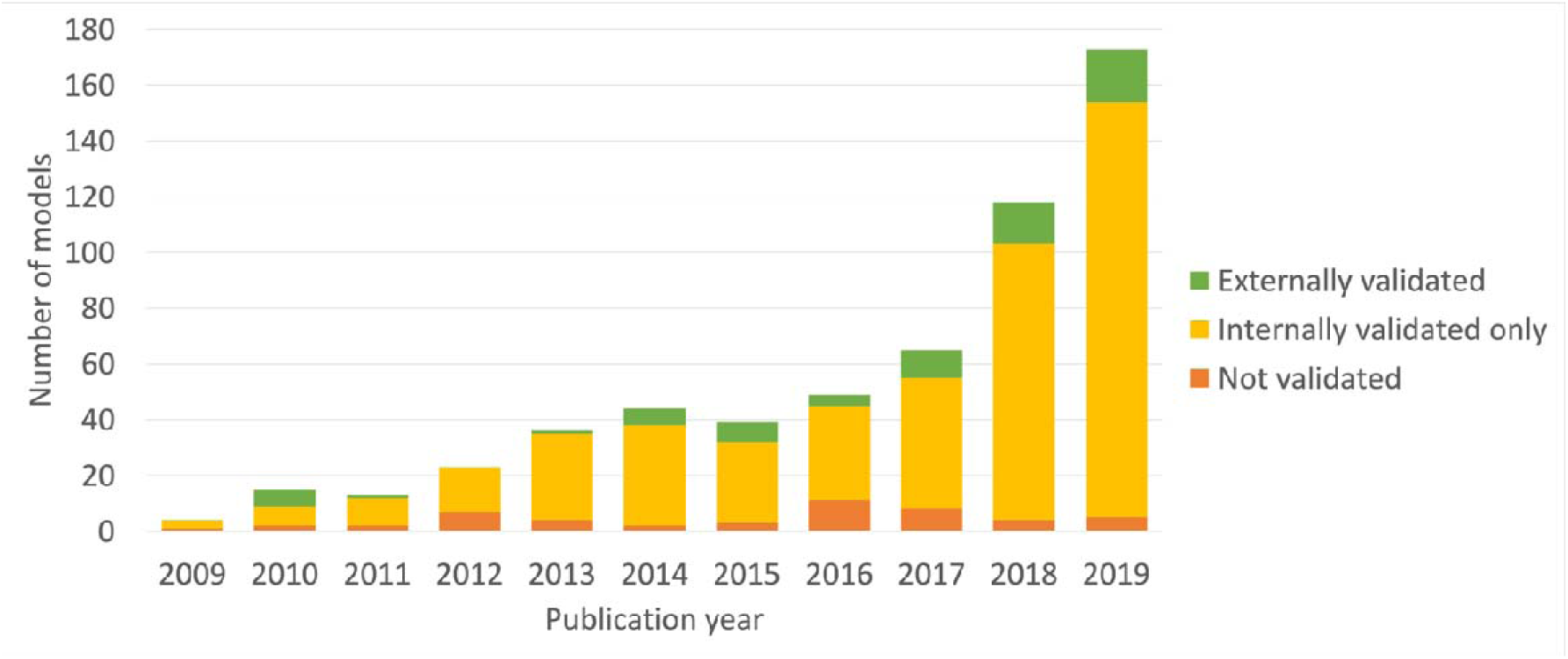
Trends in model validation

Internal validation results were reported for a total of 525 models in 382 studies (Table 4). This includes a total of 64 models in 43 studies for which both internal and external validation results were reported. In both periods, the AUROC was reported for more than 90% (107/115 and 392/410) of all internal validations, with a median reported AUROC of slightly below 0.8 in both periods. The percentage of internal validations for which the ROC curve was presented increased from 28% (32/115) to 47% (192/410). The percentage of internal validations for which the calibration plot was presented was slightly less than 30% (33/115 and 116/410) in both periods. For about 25% (29/115 and 91/410) of all internal validations, other calibration measures (such as the calibration intercept and slope) were reported.

**Table 4.** Trends in the reporting of internal validation

| Characteristic | 2009-2014 (N=115) | 2015-2019 (N=410) |
| --- | --- | --- |
| Internal validation – AUROC reported (n, %) | 107 (93%) | 392 (96%) |
| Internal validation – AUROC value (median, IQR) | 0.78 (0.73; 0.84) | 0.79 (0.72; 0.85) |
| Internal validation – ROC curve presented (n, %) | 32 (28%) | 192 (47%) |
| Internal validation – Calibration plot presented (n, %) | 33 (29%) | 116 (28%) |
| Internal validation – Other calibration measures reported (n, %) | 29 (25%) | 91 (22%) |

External validation results were reported for a total of 69 models in 45 studies. For models with multiple external validations, we focused on the results that were based on the largest reported number of observations. The number of observations in the validation dataset was reported for almost all (14/14 and 54/55) external validations, where the median reported number increased from 5,189 (IQR: 1,155; 85,048) to 27,905 (IQR: 3,189; 189,082). The number of outcome events was reported for less than 80% of the external validations in both periods (11/14 and 39/55), where the median reported number increased from 689 (IQR: 42; 1,297) to 1,014 (IQR: 247; 5,108). The AUROC was reported for almost all (13/14 and 54/55) external validations, with a median reported AUROC of slightly below 0.8 in both periods. The percentage of external validations for which the ROC curve was presented decreased from 64% (9/14) to 27% (15/55). In both periods, the calibration plot was presented for less than 30% (4/14 and 13/55) of all external validations, while other calibration measures were reported for about 15% (2/14 and 8/55) of all external validations.

## DISCUSSION

In the period 2009-2019, we found 422 papers describing the development of a total of 579 prognostic prediction models using EHR data. We observed a steep increase over the years in the number of models and an increase in the number of countries from which the EHR data that were used, originated. Overall, we observed limited improvement over time in the conduct and reporting of model development and validation. In particular, the reporting of information necessary to enable external validation by other investigators of the developed prediction models was often incomplete, with little to no improvement over time. To the best of our knowledge, no previous study has systematically evaluated these trends.

Importantly, we found that throughout the period 2009-2019, code lists to define the target population and the outcome were rarely provided, and the time-at-risk period was often unclear. Such reporting leaves other investigators guessing the exact prediction problem and how the model would translate to clinical practice; hence, the reporting of information necessary to enable external validation by other investigators was incomplete.

Most models were developed using regression analysis. For regression models, the final model presentation is usually relatively straightforward; TRIPOD recommends presenting the final model using a full model equation including all coefficients and the intercept or baseline.[8] However, for about half of the regression models throughout the period 2009-2019, the final model was not completely presented. In this way, the reporting of information necessary to enable external validation by other investigators was also incomplete.

We observed an overall increase in the use of ensemble learning and neural networks. For these modelling methods, the final model can in most cases not be presented using a full model equation such as with regression models. TRIPOD explicitly focuses on models developed using regression analysis and therefore provides limited guidance on how to present the final model for non-regression models.[8] Sharing an online tool or a software package containing the analytical source code could be a suitable alternative presentation.[6] However, we found that such resources were rarely provided. Additional guidelines for the more complex modelling methods are currently under development.[14]

Data characteristics, data handling, and validation results should always be reported to allow other investigators to interpret the findings. In line with findings from previous reviews,[1, 4, 5] we found that in the period 2009-2014, many studies made no explicit mention of how missing data were handled, model calibration was often not assessed, and external validation was uncommon. Also, these aspects barely improved over time. While the number of observations and the number of outcome events were often reported, the reporting of how missing data were handled remained at about half of the models. We did not observe an improvement in the reporting of model calibration; the calibration plot was only presented for less than one third of the models. The AUROC was reported for most models throughout the period 2009-2019, although the ROC curve itself was often not presented.

Only a small proportion of the developed models was presented along with external validation results, although with an increase over time. This further highlights the need to enable external validation by other investigators. In addition to that, very few models were externally validated using data from another country, while this would be valuable to evaluate generalizability and robustness of the prediction performance across countries.

A potential limitation of our study is that there may be eligible papers that we did not capture. In the literature, various terms have been used interchangeably to describe EHR data. Additionally, terminology surrounding prediction modelling is inconsistent.[1] However, we consulted information specialists with expert knowledge of medical terminology and databases and used a broad search to maximize the retrieval of eligible papers.

Our review clearly shows that improvement in the conduct and reporting of model development and validation is still urgently needed. Ongoing advances in the medical informatics field may aid in improving the conduct and reporting of model development and validation using EHR data: 1) improved interoperability of health data will allow researchers to validate their results more easily across centers that use different EHRs, and 2) the use of standardized analytics pipelines that enforce best practices will allow researchers to follow relevant guidelines such as TRIPOD. For example, to improve the interoperability of originally heterogeneous data sources, the Observational Health Data Sciences and Informatics (OHDSI) collaborative uses the Observational Medical Outcomes Partnership Common Data Model (OMOP CDM), which transforms source data into a common format using a set of common terminologies, vocabularies, and coding schemes. The OHDSI PatientLevelPrediction framework in turn enables a standardized analytics pipeline for the development and validation of clinical prediction models across databases that are all mapped to the OMOP CDM, while enforcing best practices based on relevant existing guidelines (including TRIPOD). [7, 15] By using such a pipeline, researchers can more easily improve the reporting of information necessary to enable external validation by other investigators.

## CONCLUSION

Before implementing a prediction model in clinical practice, it is important to ensure its prediction performance is generalizable and robust by externally validating the model across various databases. This systematic review aimed to provide further insights into the conduct and reporting of clinical prediction model development and validation over time. We focused on assessing the reporting of information necessary to enable external validation by other investigators in studies that developed models using EHR data published in the period 2009-2019. We found that the prediction problem definition was often not clearly reported, and the final model was often not completely presented, with little to no improvement over time. Thus, improvement in the reporting of information necessary to enable external validation by other investigators is still urgently needed to increase clinical adoption of developed models.

## Data Availability

The data underlying this article are available upon request.

## ACKNOWLEDGEMENTS

The authors would like to thank Christa Niehot and Dr. Wichor Bramer from the Erasmus University Medical Center medical library for their input on the search strategy. The authors would also like to thank Dr. Jenna Reps for her input on an earlier draft of this manuscript.

## DATA AVAILABILITY STATEMENT

The data underlying this article are available upon request.

## REFERENCES

1. Collins, G.S., et al., External validation of multivariable prediction models: a systematic review of methodological conduct and reporting. BMC Med Res Methodol, 2014. 14: p. 40.

2. Collins, G.S., E.O. Ogundimu, and D.G. Altman, Sample size considerations for the external validation of a multivariable prognostic model: a resampling study. Stat Med, 2016. 35(2): p. 214–26.

3. Van Calster, B., et al., Predictive analytics in health care: how can we know it works? J Am Med Inform Assoc, 2019. 26(12): p. 1651–1654.

4. Bouwmeester, W., et al., Reporting and methods in clinical prediction research: a systematic review. PLoS Med, 2012. 9(5): p. 1–12.

5. Goldstein, B.A., et al., Opportunities and challenges in developing risk prediction models with electronic health records data: a systematic review. J Am Med Inform Assoc, 2017. 24(1): p. 198–208.

6. Bonnett, L.J., et al., Guide to presenting clinical prediction models for use in clinical settings. BMJ, 2019. 365: p. l737.

7. Reps, J.M., et al., Design and implementation of a standardized framework to generate and evaluate patient-level prediction models using observational healthcare data. Journal of the American Medical Informatics Association, 2018. 25(8): p. 969–975.

8. Collins, G.S., et al., Transparent reporting of a multivariable prediction model for individual prognosis or diagnosis (TRIPOD): the TRIPOD statement. BMJ, 2015. 350: p. g7594.

9. Najafabadi, A.H.Z., et al., TRIPOD statement: a preliminary pre-post analysis of reporting and methods of prediction models. BMJ open, 2020. 10(9): p. e041537.

10. Liberati, A., et al., The PRISMA statement for reporting systematic reviews and meta-analyses of studies that evaluate health care interventions: explanation and elaboration. J Clin Epidemiol, 2009. 62(10): p. e1–34.

11. Moons, K.G., et al., Critical appraisal and data extraction for systematic reviews of prediction modelling studies: the CHARMS checklist. PLoS Med, 2014. 11(10): p. e1001744.

12. Steyerberg, E.W., et al., Assessing the performance of prediction models: a framework for some traditional and novel measures. Epidemiology (Cambridge, Mass.), 2010. 21(1): p. 128.

13. Van Calster, B., et al., Calibration: the Achilles heel of predictive analytics. BMC medicine, 2019. 17(1): p. 1–7.

14. Collins, G.S., et al., Protocol for development of a reporting guideline (TRIPOD-AI) and risk of bias tool (PROBAST-AI) for diagnostic and prognostic prediction model studies based on artificial intelligence. BMJ open, 2021. 11(7): p. e048008.

15. Khalid, S., et al., A standardized analytics pipeline for reliable and rapid development and validation of prediction models using observational health data. Computer Methods and Programs in Biomedicine, 2021. 211: p. 106394.

