## Appendix A for "Trends in the conduct and reporting of clinical prediction model development and validation: a systematic review"

### APPENDIX A. SEARCH QUERIES

#### Embase

('prediction model'/de OR 'prediction'/de OR ((predict* NEAR/3 (model* OR risk* OR rule*)):ab,kw) OR (predict* OR ((risk*) NEAR/6 (model* OR estimat* OR adjust* OR assess* OR identif* OR validat* OR forecast* OR scor*))):ti) **AND** ('medical record'/exp OR (EHR OR EHRs OR EMR OR EMRs OR ((electronic*) NEAR/3 (record* OR data*)) OR ((patient*) NEAR/3 (record* OR histor* OR data*) NEAR/6 (electronic* OR online* OR digital* OR computeri* OR automat*))):ab,ti,kw) AND [2009-2020]/py AND [english]/lim NOT ([conference abstract]/lim OR 'conference review'/it OR 'editorial'/it OR 'letter'/it OR 'note'/it OR 'review'/it OR 'tombstone'/it OR 'short survey'/it) NOT ([animals]/lim NOT [humans]/lim)

#### Medline

("Decision Support Techniques"/ OR ((predict* ADJ3 (model* OR risk* OR rule*)).ab,kw.) OR (predict* OR ((risk*) ADJ6 (model* OR estimat* OR adjust* OR assess* OR identif* OR validat* OR forecast* OR scor*))).ti.) **AND** (exp "Medical Records Systems, Computerized"/ OR (EHR OR EHRs OR EMR OR EMRs OR ((electronic*) ADJ3 (record* OR data*)) OR ((patient*) ADJ3 (record* OR histor* OR data*) ADJ6 (electronic* OR online* OR digital* OR computeri* OR automat*))).ab,ti,kw.) AND (english).lg NOT (news OR congres* OR abstract* OR book* OR chapter* OR dissertation abstract* OR editorial* OR letter*).pt. NOT (exp Animals/ NOT Humans/) AND (limit 1 to yr=2009-2020)

#### Web-of-Science

TS=((((predict* NEAR/2 (model* OR risk* OR rule*))) OR (predict* OR ((risk*) NEAR/5 (model* OR estimat* OR adjust* OR assess* OR identif* OR validat* OR forecast* OR scor*))):ti) **AND** ((EHR OR EHRs OR EMR OR EMRs OR ((electronic*) NEAR/2 (record* OR data*)) OR ((patient*) NEAR/2 (record* OR histor* OR data*) NEAR/5 (electronic* OR online* OR digital* OR computeri*)))) NOT ((animal* OR rat OR rats OR mouse OR mice OR murine OR dog OR dogs OR canine OR cat OR cats OR feline OR rabbit OR cow OR cows OR bovine OR rodent* OR sheep OR ovine OR pig OR swine OR porcine OR veterinar* OR chick* OR zebrafish* OR baboon* OR nonhuman* OR primate* OR cattle* OR goose OR geese OR duck OR macaque* OR avian* OR bird* OR fish*) NOT (human* OR patient* OR women OR woman OR men OR man))) AND PY=(2009-2020) AND LA=(English) AND DT=(Article OR Review)

#### Cochrane Library

(((predict* NEAR/3 (model* OR risk* OR rule*)).ab,kw.) OR (predict* OR ((risk*) NEAR/6 (model* OR estimat* OR adjust* OR assess* OR identif* OR validat* OR forecast* OR scor*))):ti) **AND** ((EHR OR EHRs OR EMR OR EMRs OR ((electronic*) NEAR/3 (record* OR data*)) OR ((patient*) NEAR/3 (record* OR histor* OR data*) NEAR/6 (electronic* OR online* OR digital* OR computeri*))):ab,ti,kw)

#### Google Scholar

"clinical decision model"|"clinical decision models" "electronic health record"|"electronic health data"|"electronic health records"
