## Appendix B for "Trends in the conduct and reporting of clinical prediction model development and validation: a systematic review"

### APPENDIX B. STUDY SELECTION FLOW DIAGRAM

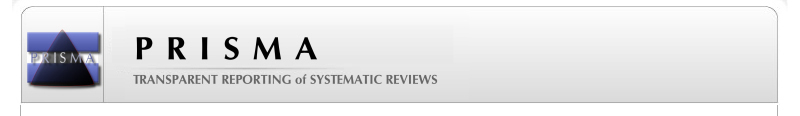
**PRISMA 2009 Flow Diagram**

Records excluded
(n = 5,160)

Full-text articles excluded
(n = 653)

- Not using EHR data (n = 159)
- Methodological focus (n = 140)
- Evaluating predictor associations (n = 95)
- Not prognostic prediction (n = 86)
- Not patient-level prediction of a particular clinical outcome (n = 77)
- Other article types (n = 41)
- Model updating study (n = 27)
- External validation study (n = 22)
- Full text unavailable (n = 6)

Studies included for data extraction
(n = 422)

Full-text articles assessed for eligibility
(n = 1,075)

Records screened
(n = 6,235)

Records after duplicates removed
(n =	6,235)

Records identified through database searching
(n = 9,942)

#### Identification

#### Eligibility

#### Included

#### Screening
