## Appendix C for "Trends in the conduct and reporting of clinical prediction model development and validation: a systematic review"

### APPENDIX C. REFERENCE LIST OF INCLUDED STUDIES

## 2009

A1. Himes, B.E., et al., Prediction of Chronic Obstructive Pulmonary Disease (COPD) in Asthma Patients Using Electronic Medical Records. J Am Med Informatics Assoc, 2009. 16(3): p. 371-379.
A2. Li, C.H., et al., Predictive Model for Length of Hospital Stay of Patients Surviving Surgery for Perforated Peptic Ulcer. J Formos Med Assoc, 2009. 108(8): p. 644-652.
A3. Reis, B.Y., I.S. Kohane, and K.D. Mandl, Longitudinal histories as predictors of future diagnoses of domestic abuse: modelling study. BMJ, 2009. 339: p. b3677.
A4. Smits, F.T.M., et al., Predictability of persistent frequent attendance: A historic 3-year cohort study. Br J Gen Pract, 2009. 59(559): p. 114-119.

## 2010

A5. Amarasingham, R., et al., An automated model to identify heart failure patients at risk for 30-day readmission or death using electronic medical record data. Med Care, 2010. 48(11): p. 981-988.
A6. Calvillo-King, L., et al., Predicting risk of perioperative death and stroke after carotid endarterectomy in asymptomatic patients: Derivation and validation of a clinical risk score. Stroke, 2010. 41(12): p. 2786-2794.
A7. Crane, S.J., et al., Use of an electronic administrative database to identify older community dwelling adults at high-risk for hospitalization or emergency department visits: the elders risk assessment index. BMC Health Serv Res, 2010. 10: p. 338.
A8. Daggy, J., et al., Using no-show modeling to improve clinic performance. Health Informatics Journal, 2010. 16(4): p. 246-259.
A9. Graesslin, O., et al., Nomogram to predict subsequent brain metastasis in patients with metastatic breast cancer. J Clin Oncol, 2010. 28(12): p. 2032-2037.
A10. Johnson, E.S., et al., Predicting the risk of hyperkalemia in patients with chronic kidney disease starting lisinopril. Pharmacoepidemiol Drug Saf, 2010. 19(3): p. 266-272.
A11. Matheny, M.E., et al., Development of inpatient risk stratification models of acute kidney injury for use in electronic health records. Med Decis Making, 2010. 30(6): p. 639-650.
A12. Robbins, G.K., et al., Predicting virologic failure in an HIV clinic. Clin Infect Dis, 2010. 50(5): p. 779-786.
A13. Sessler, D.I., et al., Broadly applicable risk stratification system for predicting duration of hospitalization and mortality. Anesthesiology, 2010. 113(5): p. 1026-1037.
A14. Tude Melo, J.R., et al., Mortality in children with severe head trauma: Predictive factors and proposal for a new predictive scale. Neurosurgery, 2010. 67(6): p. 1542-1547.
A15. Wu, J., J. Roy, and W.F. Stewart, Prediction modeling using EHR data: Challenges, strategies, and a comparison of machine learning approaches. Med Care, 2010. 48(6 SUPPL.): p. S106-S113.

## 2011

A16. Barrett, T.W., et al., A clinical prediction model to estimate risk for 30-day adverse events in emergency department patients with symptomatic atrial fibrillation. Ann Emerg Med, 2011. 57(1): p. 1-12.
A17. Braga, M., et al., A prognostic score to predict major complications after pancreaticoduodenectomy. Ann Surg, 2011. 254(5): p. 702-707.
A18. Chang, Y.J., et al., Predicting hospital-acquired infections by scoring system with simple parameters. PLoS ONE, 2011. 6(8).
A19. Cheng, P., et al., Hemoglobin A1c as a predictor of incident diabetes. Diabetes Care, 2011. 34(3): p. 610-615.
A20. Dubberke, E.R., et al., Development and validation of a Clostridium difficile infection risk prediction model. Infect Control Hosp Epidemiol, 2011. 32(4): p. 360-366.
A21. Kor, D.J., et al., Derivation and diagnostic accuracy of the surgical lung injury prediction model. Anesthesiology, 2011. 115(1): p. 117-128.
A22. Meyfroidt, G., et al., Computerized prediction of intensive care unit discharge after cardiac surgery: development and validation of a Gaussian processes model. BMC Med Inform Decis Mak, 2011. 11: p. 64.
A23. Smith, D.H., et al., Predicting poor outcomes in heart failure. Perm. j., 2011. 15(4): p. 4-11.
A24. Watson, A.J., et al., Linking Electronic Health Record-Extracted Psychosocial Data in Real-Time to Risk of Readmission for Heart Failure. Psychosomatics, 2011. 52(4): p. 319-327.
A25. Westra, B.L., et al., Predicting improvement in urinary and bowel incontinence for home health patients using electronic health record data. J Wound Ostomy Continence Nurs, 2011. 38(1): p. 77-87.
A26. Woller, S.C., et al., Derivation and validation of a simple model to identify venous thromboembolism risk in medical patients. Am J Med, 2011. 124(10): p. 947-954.e2.
A27. Zhao, D. and C. Weng, Combining PubMed knowledge and EHR data to develop a weighted bayesian network for pancreatic cancer prediction. J Biomed Informatics, 2011. 44(5): p. 859-868.

## 2012

A28. Casarett, D.J., et al., The art versus science of predicting prognosis: Can a prognostic index predict short-term mortality better than experienced nurses do? J Palliative Med, 2012. 15(6): p. 703-708.
A29. Elliott, M.B., et al., Prediction and prevention of treatment-related inpatient hypoglycemia. J Diabetes Sci Technol, 2012. 6(2): p. 302-309.
A30. Escobar, G.J., et al., Early detection of impending physiologic deterioration among patients who are not in intensive care: Development of predictive models using data from an automated electronic medical record. J Hosp Med, 2012. 7(5): p. 388-395.
A31. Hsu, C.C., et al., Early Mortality Risk Score: Identification of Poor Outcomes Following Upfront Surgery for Resectable Pancreatic Cancer. J Gastrointest Surg, 2012. 16(4): p. 753-761.
A32. Jonikas, M.A. and K.D. Mandl, Surveillance of medication use: early identification of poor adherence. J Am Med Inform Assoc, 2012. 19(4): p. 649-654.
A33. Karnik, S., et al., Predicting atrial fibrillation and flutter using electronic health records. Conf Proc IEEE Eng Med Biol Soc, 2012. 2012: p. 5562-5565.
A34. Kawaler, E., et al., Learning to predict post-hospitalization VTE risk from EHR data. AMIA Annu Symp Proc, 2012. 2012: p. 436-445.
A35. Khan, A., et al., An electronic medical record-derived real-time assessment scale for hospital readmission in the elderly. Wisc Med J, 2012. 111(3): p. 119-122.
A36. Kramer, D.B., et al., Development and validation of a risk score to predict early mortality in recipients of implantable cardioverter-defibrillators. Heart Rhythm, 2012. 9(1): p. 42-46.
A37. Levin, S.R., et al., Real-time forecasting of pediatric intensive care unit length of stay using computerized provider orders. Crit Care Med, 2012. 40(11): p. 3058-3064.
A38. Mani, S., et al., Type 2 diabetes risk forecasting from EMR data using machine learning. AMIA Annu Symp Proc, 2012. 2012: p. 606-615.
A39. Monsen, K.A., et al., Exploring the value of clinical data standards to predict hospitalization of home care patients. Appl Clin Informatics, 2012. 3(4): p. 419-436.
A40. Navarro, A.E., S. Enguídanos, and K.H. Wilber, Identifying risk of hospital readmission among medicare aged patients: An approach using routinely collected data. Home Health Care Serv Q, 2012. 31(2): p. 181-195.
A41. Nijhawan, A.E., et al., An electronic medical record-based model to predict 30-day risk of readmission and death among HIV-infected inpatients. J Acquired Immune Defic Syndr, 2012. 61(3): p. 349-358.
A42. Tescher, A.N., et al., All at-risk patients are not created equal: Analysis of braden pressure ulcer risk scores to identify specific risks. J Wound Ostomy Continence Nurs, 2012. 39(3): p. 282-291.
A43. Wang, L., et al., Predicting risk of hospitalization or death among patients with heart failure in the veterans health administration. Am J Cardiol, 2012. 110(9): p. 1342-1349.

## 2013

A44. Alvarez, C.A., et al., Predicting out of intensive care unit cardiopulmonary arrest or death using electronic medical record data. BMC Med Inform Decis Mak, 2013. 13: p. 28.
A45. Baillie, C.A., et al., The readmission risk flag: Using the electronic health record to automatically identify patients at risk for 30-day readmission. J Hosp Med, 2013. 8(12): p. 689-695.
A46. Billings, J., et al., Choosing a model to predict hospital admission: An observational study of new variants of predictive models for case finding. BMJ Open, 2013. 3(8).
A47. Brueckmann, B., et al., Development and validation of a score for prediction of postoperative respiratory complications. Anesthesiology, 2013. 118(6): p. 1276-1285.
A48. Choudhry, S.A., et al., A public-private partnership develops and externally validates a 30-day hospital readmission risk prediction model. Online j. public health inform., 2013. 5(2): p. 219.
A49. Conrad, M.F., et al., A risk prediction model for determining appropriateness of CEA in patients with asymptomatic carotid artery stenosis. Ann Surg, 2013. 258(4): p. 534-538.
A50. Cooper, P.B., A.J. Heuer, and C.A. Warren, Electronic screening of patients for predisposition to Clostridium difficile infection in a community hospital. Am J Infect Control, 2013. 41(3): p. 232-235.
A51. Eapen, Z.J., et al., Validated, electronic health record deployable prediction models for assessing patient risk of 30-day rehospitalization and mortality in older heart failure patients. JACC Heart Fail, 2013. 1(3): p. 245-251.
A52. Escobar, G.J., et al., Risk-adjusting hospital mortality using a comprehensive electronic record in an integrated health care delivery system. Med Care, 2013. 51(5): p. 446-453.
A53. Giunta, D., et al., Factors associated with nonattendance at clinical medicine scheduled outpatient appointments in a university general hospital. Patient Preference Adherence, 2013. 7: p. 1163-1170.
A54. Hebert, C., et al., Electronic health record-based detection of risk factors for clostridium difficile infection relapse. Infect Control Hosp Epidemiol, 2013. 34(4): p. 407-414.
A55. Herzig, S.J., et al., Risk factors for nosocomial gastrointestinal bleeding and use of acid-suppressive medication in non-critically ill patients. J Gen Intern Med, 2013. 28(5): p. 683-690.
A56. Jin, S.J., et al., A new statistical approach to predict bacteremia using electronic medical records. Scand J Infect Dis, 2013. 45(9): p. 672-680.
A57. Kennedy, E.H., et al., Improved cardiovascular risk prediction using nonparametric regression and electronic health record data. Med Care, 2013. 51(3): p. 251-258.
A58. Kim, C., et al., Prediction of metachronous multiple primary cancers following the curative resection of gastric cancer. BMC Cancer, 2013. 13.
A59. Mathias, J.S., et al., Development of a 5 year life expectancy index in older adults using predictive mining of electronic health record data. J Am Med Informatics Assoc, 2013. 20(E1): p. e118-e124.
A60. Ramchandran, K.J., et al., A predictive model to identify hospitalized cancer patients at risk for 30-day mortality based on admission criteria via the electronic medical record. Cancer, 2013. 119(11): p. 2074-2080.
A61. Revel-Vilk, S., et al., Age and duration of bleeding symptoms at diagnosis best predict resolution of childhood immune thrombocytopenia at 3, 6, and 12 months. J Pediatr, 2013. 163(5): p. 1335-1339.e2.
A62. Singal, A.G., et al., An automated model using electronic medical record data identifies patients with cirrhosis at high risk for readmission. Clin Gastroenterol Hepatol, 2013. 11(10): p. 1335-1341.
A63. Tabak, Y.P., et al., Using enriched observational data to develop and validate age-specific mortality risk adjustment models for hospitalized pediatric patients. Med Care, 2013. 51(5): p. 437-445.
A64. Tabak, Y.P., et al., Development and Validation of a Mortality Risk-Adjustment Model for Patients Hospitalized for Exacerbations of Chronic Obstructive Pulmonary Disease. Med Care, 2013. 51(7): p. 597-605.
A65. Wang, L., et al., Predicting risk of hospitalization or death among patients receiving primary care in the veterans health administration. Med Care, 2013. 51(4): p. 368-373.
A66. Wells, B.J., et al., Prediction of morbidity and mortality in patients with type 2 diabetes. PeerJ, 2013. 2013(1).
A67. Wong, R.M., C. Bresee, and G.D. Braunstein, Comparison with published systems of a new staging system for papillary and follicular thyroid carcinoma. Thyroid, 2013. 23(5): p. 566-574.

## 2014

A68. Atchison, C.M., et al., Development of a new risk score for hospital-associated venous thromboembolism in noncritically ill children: Findings from a large single-institutional case-control study. J Pediatr, 2014. 165(4): p. 793-798.
A69. Ayyagari, R., et al., Pulse pressure and stroke risk: development and validation of a new stroke risk model. Curr Med Res Opin, 2014. 30(12): p. 2453-2460.
A70. Bayati, M., et al., Data-driven decisions for reducing readmissions for heart failure: General methodology and case study. PLoS ONE, 2014. 9(10).
A71. Carter, E.M. and H.W. Potts, Predicting length of stay from an electronic patient record system: a primary total knee replacement example. BMC Med Inform Decis Mak, 2014. 14: p. 26.
A72. Castro, V.M., et al., Stratification of risk for hospital admissions for injury related to fall: cohort study. Bmj, 2014. 349: p. g5863.
A73. Chase, H.S., et al., Presence of early CKD-related metabolic complications predict progression of stage 3 CKD: A case-controlled study. BMC Nephrol, 2014. 15(1).
A74. Churpek, M.M., et al., Using electronic health record data to develop and validate a prediction model for adverse outcomes in the wards. Crit Care Med, 2014. 42(4): p. 841-848.
A75. Churpek, M.M., et al., Multicenter development and validation of a risk stratification tool for ward patients. Am J Respir Crit Care Med, 2014. 190(6): p. 649-655.
A76. Cobellis, L., et al., Is it possible to predict office hysteroscopy failure? 2014. 181: p. 328-333.
A77. Goldstein, B.A., et al., Near-term prediction of sudden cardiac death in older hemodialysis patients using electronic health records. Clin J Am Soc Nephrol, 2014. 9(1): p. 82-91.
A78. Gong, C.S.A., et al., Predicting postoperative vomiting for orthopedic patients receiving patient-controlled epidural analgesia with the application of an artificial neural network. BioMed Res Int, 2014. 2014.
A79. Gultepe, E., et al., From vital signs to clinical outcomes for patients with sepsis: A machine learning basis for a clinical decision support system. J Am Med Informatics Assoc, 2014. 21(2): p. 315-325.
A80. Gupta, S., et al., Machine-learning prediction of cancer survival: A retrospective study using electronic administrative records and a cancer registry. BMJ Open, 2014. 4(3).
A81. Hao, S., et al., Risk prediction of emergency department revisit 30 days post discharge: A prospective study. PLoS ONE, 2014. 9(11).
A82. He, D., et al., Mining high-dimensional administrative claims data to predict early hospital readmissions. J Am Med Inform Assoc, 2014. 21(2): p. 272-279.
A83. Hebert, C., et al., Diagnosis-specific readmission risk prediction using electronic health data: a retrospective cohort study. BMC Med Inform Decis Mak, 2014. 14: p. 65.
A84. Huang, S.H., et al., Toward personalizing treatment for depression: Predicting diagnosis and severity. J Am Med Informatics Assoc, 2014. 21(6): p. 1069-1075.
A85. Huang, Y. and D.A. Hanauer, Patient no-show predictive model development using multiple data sources for an effective overbooking approach. Appl Clin Inform, 2014. 5(3): p. 836-860.
A86. Je, H.U., et al., A nomogram predicting the risks of distant metastasis following postoperative radiotherapy for uterine cervical carcinoma: A Korean radiation oncology group study (KROG 12-08). Radiother Oncol, 2014. 111(3): p. 437-441.
A87. Kim, S.H., et al., Postoperative nomogram to predict the probability of metastasis in enneking stage IIB extremity osteosarcoma. BMC Cancer, 2014. 14(1).
A88. Kontio, E., et al., Predicting patient acuity from electronic patient records. J Biomed Informatics, 2014. 51: p. 35-40.
A89. O'Leary, E.A., et al., Letting the sun set on small bowel obstruction: Can a simple risk score tell us when nonoperative care is inappropriate? Am Surg, 2014. 80(6): p. 572-579.
A90. Puttkammer, N., et al., Development of an electronic medical record based alert for risk of HIV treatment failure in a low-resource setting. PLoS ONE, 2014. 9(11).
A91. Rana, S., et al., Predicting unplanned readmission after myocardial infarction from routinely collected administrative hospital data. Aust Health Rev, 2014. 38(4): p. 377-382.
A92. Rapsomaniki, E., et al., Prognostic models for stable coronary artery disease based on electronic health record cohort of 102 023 patients. Eur Heart J, 2014. 35(13): p. 844-852.
A93. Roubinian, N.H., et al., Predicting red blood cell transfusion in hospitalized patients: role of hemoglobin level, comorbidities, and illness severity. BMC Health Serv Res, 2014. 14: p. 213.
A94. Sho, S., et al., A novel scoring system to predict the development of necrotizing enterocolitis totalis in premature infants. J Pediatr Surg, 2014. 49(7): p. 1053-1056.
A95. Still, C.D., et al., Preoperative prediction of type 2 diabetes remission after Roux-en-Y gastric bypass surgery: A retrospective cohort study. Lancet Diabetes Endocrinol, 2014. 2(1): p. 38-45.
A96. Sun, J., et al., Predicting changes in hypertension control using electronic health records from a chronic disease management program. J Am Med Informatics Assoc, 2014. 21(2): p. 337-344.
A97. Tabak, Y.P., et al., Using electronic health record data to develop inpatient mortality predictive model: Acute Laboratory Risk of Mortality Score (ALaRMS). J Am Med Informatics Assoc, 2014. 21(3): p. 455-463.
A98. Taha, M., et al., Derivation and validation of a formula to estimate risk for 30-day readmission in medical patients. Int J Qual Health Care, 2014. 26(3): p. 271-277.
A99. Tran, T., et al., Risk stratification using data from electronic medical records better predicts suicide risks than clinician assessments. BMC Psychiatry, 2014. 14(1).
A100. Wiens, J., et al., Learning data-driven patient risk stratification models for Clostridium difficile. Open Forum Infect Dis, 2014. 1(2).
A101. Zhai, H., et al., Developing and evaluating a machine learning based algorithm to predict the need of pediatric intensive care unit transfer for newly hospitalized children. Resuscitation, 2014. 85(8): p. 1065-1071.

## 2015

A102. Amarasingham, R., et al., Electronic medical record-based multicondition models to predict the risk of 30 day readmission or death among adult medicine patients: validation and comparison to existing models. BMC Med Inform Decis Mak, 2015. 15: p. 39.
A103. Blumenthal, D.M., et al., Predicting Non-Adherence with Outpatient Colonoscopy Using a Novel Electronic Tool that Measures Prior Non-Adherence. J Gen Intern Med, 2015. 30(6): p. 724-731.
A104. Brown, J.R., et al., Acute kidney injury risk prediction in patients undergoing coronary angiography in a national veterans health administration cohort with external validation. J Am Heart Assoc, 2015. 4(12).
A105. Chan, S., et al., Prediction of intracerebral haemorrhage expansion with clinical, laboratory, pharmacologic, and noncontrast radiographic variables. Int J Stroke, 2015. 10(7): p. 1057-1061.
A106. Cronin, R.M., et al., National Veterans Health Administration inpatient risk stratification models for hospital-acquired acute kidney injury. J Am Med Informatics Assoc, 2015. 22(5): p. 1054-1071.
A107. Dai, W., et al., Prediction of hospitalization due to heart diseases by supervised learning methods. Int J Med Informatics, 2015. 84(3): p. 189-197.
A108. Eby, E., et al., Predictors of 30 day hospital readmission in patients with type 2 diabetes: A retrospective, case-control, database study. Curr Med Res Opin, 2015. 31(1): p. 107-114.
A109. Escobar, G.J., et al., Nonelective rehospitalizations and postdischarge mortality predictive models suitable for use in real time. Med Care, 2015. 53(11): p. 916-923.
A110. Hao, S., et al., Development, Validation and Deployment of a Real Time 30 Day Hospital Readmission Risk Assessment Tool in the Maine Healthcare Information Exchange. PLoS ONE, 2015. 10(10): p. e0140271.
A111. Hippisley-Cox, J. and C. Coupland, Development and validation of risk prediction equations to estimate future risk of heart failure in patients with diabetes: A prospective cohort study. BMJ Open, 2015. 5(9).
A112. Hu, Z., et al., Real-time web-based assessment of total population risk of future emergency department utilization: statewide prospective active case finding study. Interact J Med Res, 2015. 4(1): p. e2.
A113. Hylan, T.R., et al., Automated prediction of risk for problem opioid use in a primary care setting. J Pain, 2015. 16(4): p. 380-387.
A114. Kaewprag, P., et al., Predictive Modeling for Pressure Ulcers from Intensive Care Unit Electronic Health Records. AMIA Summits Transl Sci Proc, 2015. 2015: p. 82-86.
A115. Montserrat-Capdevila, J., et al., Predictive model of hospital admission for COPD exacerbation. Respir Care, 2015. 60(9): p. 1288-1294.
A116. Montserrat-Capdevila, J., et al., Risk of exacerbation in chronic obstructive pulmonary disease: a primary care retrospective cohort study. BMC Fam Pract, 2015. 16: p. 173.
A117. Osborn, D.P., et al., Cardiovascular risk prediction models for people with severe mental illness: results from the prediction and management of cardiovascular risk in people with severe mental illnesses (PRIMROSE) research program. JAMA Psychiatry, 2015. 72(2): p. 143-151.
A118. Pannu, S.R., et al., Development and validation of severe hypoxemia associated risk prediction model in 1,000 mechanically ventilated patients*. Crit Care Med, 2015. 43(2): p. 308-317.
A119. Razavian, N., et al., Population-Level Prediction of Type 2 Diabetes From Claims Data and Analysis of Risk Factors. Big data, 2015. 3(4): p. 277-287.
A120. Shadmi, E., et al., Predicting 30-day readmissions with preadmission electronic health record data. Med Care, 2015. 53(3): p. 283-289.
A121. Smolin, B., et al., Predicting mortality of elderly patients acutely admitted to the Department of Internal Medicine. Int J Clin Pract, 2015. 69(4): p. 501-508.
A122. Tabak, Y.P., et al., Predicting the Risk for Hospital-Onset Clostridium difficile Infection (HO-CDI) at the Time of Inpatient Admission: HO-CDI Risk Score. Infect Control Hosp Epidemiol, 2015. 36(6): p. 695-701.
A123. Temple, M.W., C.U. Lehmann, and D. Fabbri, Predicting discharge dates from the nicu using progress note data. Pediatrics, 2015. 136(2): p. e395-e405.
A124. Tighe, P.J., et al., Teaching a Machine to Feel Postoperative Pain: Combining High-Dimensional Clinical Data with Machine Learning Algorithms to Forecast Acute Postoperative Pain. Pain Med, 2015. 16(7): p. 1386-1401.
A125. Tolcher, M.C., et al., Predicting cesarean delivery after induction of labor among nulliparouswomen at term. Obstet Gynecol, 2015. 126(5): p. 1059-1068.
A126. Uyar, A., A. Bener, and H.N. Ciray, Predictive Modeling of Implantation Outcome in an In Vitro Fertilization Setting: An Application of Machine Learning Methods. Med Decis Making, 2015. 35(6): p. 714-725.
A127. Van Mourik, M.S.M., et al., Severity of Disease Estimation and Risk-Adjustment for Comparison of Outcomes in Mechanically Ventilated Patients Using Electronic Routine Care Data. Infect Control Hosp Epidemiol, 2015. 36(7): p. 807-815.
A128. Wu, S., et al., Prognostic nomogram for patients with nasopharyngeal carcinoma after intensity-modulated radiotherapy. PLoS ONE, 2015. 10(8).

## 2016

A129. Agrawal, D., et al., Predicting patients at risk for 3-day postdischarge readmissions, ED Visits, and Deaths. Med Care, 2016. 54(11): p. 1017-1023.
A130. Anderson, J.P., et al., Reverse Engineering and Evaluation of Prediction Models for Progression to Type 2 Diabetes: An Application of Machine Learning Using Electronic Health Records. J Diabetes Sci Technol, 2016. 10(1): p. 6-18.
A131. Babazade, R., et al., A nomogram for predicting the need for sciatic nerve block after total knee arthroplasty. J Anesth, 2016. 30(5): p. 864-872.
A132. Back, J.S., et al., Development and Validation of an Automated Sepsis Risk Assessment System. Res Nurs Health, 2016. 39(5): p. 317-327.
A133. Buci, S. and A. Kukeli, Survival probability in patients with liver trauma. Int J Health Care Qual Assur, 2016. 29(7): p. 778-785.
A134. Calvert, J., et al., Using electronic health record collected clinical variables to predict medical intensive care unit mortality. Ann Med Surg, 2016. 11: p. 52-57.
A135. Castro, V.M., et al., Stratifying Risk for Renal Insufficiency among Lithium-Treated Patients: An Electronic Health Record Study. Neuropsychopharmacology, 2016. 41(4): p. 1138-1143.
A136. Churpek, M.M., et al., Multicenter Comparison of Machine Learning Methods and Conventional Regression for Predicting Clinical Deterioration on the Wards. Crit Care Med, 2016. 44(2): p. 368-374.
A137. Gaskin, G.L., et al., Predictive modeling of risk factors and complications of cataract surgery. Eur J Ophthalmol, 2016. 26(4): p. 328-337.
A138. Gomes, D.G., et al., Simple risk stratification score for prognosis of syncope. J Intervent Card Electrophysiol, 2016. 47(2): p. 153-161.
A139. Hu, S.B., et al., Prediction of clinical deterioration in hospitalized adult patients with hematologic malignancies using a neural network model. PLoS ONE, 2016. 11(8).
A140. Iino, C., et al., Evaluation of scoring models for identifying the need for therapeutic intervention of upper gastrointestinal bleeding: A new prediction score model for Japanese patients. Dig Endosc, 2016. 28(7): p. 714-721.
A141. Jacobs, M.B., et al., Predictors of treatment failure in young patients undergoing in vitro fertilization. J Assisted Reprod Genet, 2016. 33(8): p. 1001-1007.
A142. Jin, B., et al., Prospective stratification of patients at risk for emergency department revisit: Resource utilization and population management strategy implications. BMC Emerg Med, 2016. 16(1).
A143. Karmakar, C., et al., Predicting Risk of Suicide Attempt Using History of Physical Illnesses From Electronic Medical Records. JMIR Ment Health, 2016. 3(3): p. e19.
A144. Kipnis, P., et al., Development and validation of an electronic medical record-based alert score for detection of inpatient deterioration outside the ICU. J Biomed Informatics, 2016. 64: p. 10-19.
A145. Koyner, J.L., et al., Development of a multicenter ward–based AKI prediction model. Clin J Am Soc Nephrol, 2016. 11(11): p. 1935-1943.
A146. Kurasawa, H., et al., Machine-Learning-Based Prediction of a Missed Scheduled Clinical Appointment by Patients with Diabetes. J Diabetes Sci Technol, 2016. 10(3): p. 730-736.
A147. Lee, J.Y., et al., Development and evaluation of an automated fall risk assessment system. Int J Qual Health Care, 2016. 28(2): p. 175-182.
A148. Logue, E., W. Smucker, and C. Regan, Admission data predict high hospital readmission risk. J Am Board Fam Med, 2016. 29(1): p. 50-59.
A149. Low, L.L., et al., Predicting 30-day readmissions in an Asian population: Building a predictive model by incorporating markers of hospitalization severity. PLoS ONE, 2016. 11(12).
A150. Low, L.L., et al., Predicting frequent hospital admission risk in Singapore: A retrospective cohort study to investigate the impact of comorbidities, acute illness burden and social determinants of health. BMJ Open, 2016. 6(10).
A151. Nguyen, O.K., et al., Predicting all-cause readmissions using electronic health record data from the entire hospitalization: Model development and comparison. J Hosp Med, 2016. 11(7): p. 473-480.
A152. Press, A., et al., Developing a Clinical Prediction Rule for First Hospital-Onset Clostridium difficile Infections: A Retrospective Observational Study. Infect Control Hosp Epidemiol, 2016. 37(8): p. 896-900.
A153. Qu, Z., et al., Building a patient-specific risk score with a large database of discharge summary reports. Med Sci Monit, 2016. 22: p. 2097-2104.
A154. Rohr, J.K., et al., Developing a predictive risk model for first-line antiretroviral therapy failure in South Africa. J Int AIDS Soc, 2016. 19(1).
A155. Rubin, D.J., et al., Developement and validation of a novel tool to predict hospital readmission resk among patients with diabetes. Endocr Pract, 2016. 22(10): p. 1204-1215.
A156. Sanchez-Pinto, L.N. and R.G. Khemani, Development of a Prediction Model of Early Acute Kidney Injury in Critically Ill Children Using Electronic Health Record Data. Pediatr Crit Care Med, 2016. 17(6): p. 508-515.
A157. Shameer, K., et al., PREDICTIVE MODELING OF HOSPITAL READMISSION RATES USING ELECTRONIC MEDICAL RECORD-WIDE MACHINE LEARNING: A CASE-STUDY USING MOUNT SINAI HEART FAILURE COHORT. Pac Symp Biocomput, 2016. 22: p. 276-287.
A158. Shen, Z., et al., Risk factors predictive of recurrence and progression for patients who suffered initial recurrence after transurethral resection of stage pT1 bladder tumor in Chinese population: A retrospective study. Medicine, 2016. 95(5).
A159. Srinivasan, A., et al., Premorbid function, comorbidity, and frailty predict outcomes after ruptured abdominal aortic aneurysm repair. J Vasc Surg, 2016. 63(3): p. 603-609.
A160. Stites, S.D., et al., The tipping point: patients predisposed to Clostridium difficile infection and a hospital antimicrobial stewardship programme. J Hosp Infect, 2016. 94(3): p. 242-248.
A161. Taylor, R.A., et al., Prediction of In-hospital Mortality in Emergency Department Patients with Sepsis: A Local Big Data-Driven, Machine Learning Approach. Acad Emerg Med, 2016. 23(3): p. 269-278.
A162. Toerper, M.F., et al., Cardiac catheterization laboratory inpatient forecast tool: A prospective evaluation. J Am Med Informatics Assoc, 2016. 23(e1): p. e49-e57.
A163. Tulloch, A.D., A.S. David, and G. Thornicroft, Exploring the predictors of early readmission to psychiatric hospital. Epidemiol Psychiatr Sci, 2016. 25(2): p. 181-193.
A164. Warren, M.B., et al., Prolonged length of stay in ED psychiatric patients: A multivariable predictive model. Am J Emerg Med, 2016. 34(2): p. 133-139.
A165. Wise, E.S., K.M. Hocking, and S.M. Kavic, Prediction of excess weight loss after laparoscopic Roux-en-Y gastric bypass: data from an artificial neural network. 2016. 30(2): p. 480-488.
A166. Wu, J., et al., A practical method for predicting frequent use of emergency department care using routinely available electronic registration data. BMC Emerg Med, 2016. 16(1).
A167. Xie, Y., et al., Risk prediction to inform surveillance of chronic kidney disease in the US Healthcare Safety Net: A cohort study. BMC Nephrol, 2016. 17(1).
A168. Yokota, S. and K. Ohe, Construction and evaluation of FiND, a fall risk prediction model of inpatients from nursing data. Jpn J Nurs Sci, 2016. 13(2): p. 247-255.
A169. Zhang, W.Y., et al., A simple scoring system predicting the survival time of patients with bone metastases after RT. PLoS ONE, 2016. 11(7).

## 2017

A170. Babu, R., et al., A new score to predict recipient mortality from preoperative donor and recipient characteristics in living donor liver transplantation (DORMAT score). Ann Transplant, 2017. 22: p. 499-506.
A171. Barak-Corren, Y., et al., Predicting suicidal behavior from longitudinal electronic health records. Am J Psychiatry, 2017. 174(2): p. 154-162.
A172. Barak-Corren, Y., A.M. Fine, and B.Y. Reis, Early prediction model of patient hospitalization from the pediatric emergency department. Pediatrics, 2017. 139(5).
A173. Barak-Corren, Y., S.H. Israelit, and B.Y. Reis, Progressive prediction of hospitalisation in the emergency department: Uncovering hidden patterns to improve patient flow. Emerg Med J, 2017. 34(5): p. 308-314.
A174. Basta, M.N., et al., Reliable prediction of postmastectomy lymphedema: The Risk Assessment Tool Evaluating Lymphedema. Am J Surg, 2017. 213(6): p. 1125-1133.e1.
A175. Baus, A., et al., An Electronic Health Record Data-driven Model for Identifying Older Adults at Risk of Unintentional Falls. Perspect Health Inf Manag, 2017. 14(Fall): p. 1b.
A176. Blakey, J.D., et al., Identifying Risk of Future Asthma Attacks Using UK Medical Record Data: A Respiratory Effectiveness Group Initiative. J Allergy Clin Immunol Pract, 2017. 5(4): p. 1015-1024.e8.
A177. Bradford, C., et al., Patient and clinical characteristics that heighten risk for heart failure readmission. Res Social Adm Pharm, 2017. 13(6): p. 1070-1081.
A178. Chen, M., et al., Development and validation of a mortality risk model for pediatric sepsis. Medicine, 2017. 96(20).
A179. Daley, M.F., et al., Predicting Hypertension Among Children With Incident Elevated Blood Pressure. Acad Pediatr, 2017. 17(3): p. 275-282.
A180. Das, L.T., et al., Predicting frequent emergency department visits among children with asthma using EHR data. Pediatr Pulmonol, 2017. 52(7): p. 880-890.
A181. Davoudi, A., et al., Delirium Prediction using Machine Learning Models on Preoperative Electronic Health Records Data. Proc IEEE Int Symp Bioinformatics Bioeng, 2017. 2017: p. 568-573.
A182. Desautels, T., et al., Prediction of early unplanned intensive care unit readmission in a UK tertiary care hospital: A cross-sectional machine learning approach. BMJ Open, 2017. 7(9).
A183. Escobar, G.J., et al., Prediction of recurrent clostridium difficile infection using comprehensive electronic medical records in an integrated healthcare delivery system. Infect Control Hosp Epidemiol, 2017. 38(10): p. 1196-1203.
A184. Frost, D.W., et al., Using the Electronic Medical Record to Identify Patients at High Risk for Frequent Emergency Department Visits and High System Costs. Am J Med, 2017. 130(5): p. 601.e17-601.e22.
A185. Fuchs, H.F., et al., Simple preoperative risk scale accurately predicts perioperative mortality following esophagectomy for malignancy. Dis Esophagus, 2017. 30(1): p. 1-6.
A186. Goshen, R., et al., Predicting the presence of colon cancer in members of a health maintenance organisation by evaluating analytes from standard laboratory records. Br J Cancer, 2017. 116(7): p. 944-950.
A187. Greenwald, J.L., et al., A Novel Model for Predicting Rehospitalization Risk Incorporating Physical Function, Cognitive Status, and Psychosocial Support Using Natural Language Processing. Med Care, 2017. 55(3): p. 261-266.
A188. Guevara, J.H., A. Zorrilla-Vaca, and G.C. Silva-Gordillo, The utility of preoperative level of erythrocytosis in the prediction of postoperative blood loss and 30-day mortality in patients with tetralogy of fallot. Ann Card Anaesth, 2017. 20(2): p. 188-192.
A189. Hao, S., et al., Estimating One-Year Risk of Incident Chronic Kidney Disease: Retrospective Development and Validation Study Using Electronic Medical Record Data From the State of Maine. JMIR Med Inform, 2017. 5(3): p. e21.
A190. Harvey, H.B., et al., Predicting No-Shows in Radiology Using Regression Modeling of Data Available in the Electronic Medical Record. J Am Coll Radiol, 2017. 14(10): p. 1303-1309.
A191. Hirji, S.A., et al., Predicting risk of cardiac events among ST-segment elevation myocardial infarction patients with conservatively managed non–infarct-related artery coronary artery disease: An analysis of the Duke Databank for Cardiovascular Disease. Am Heart J, 2017. 194: p. 116-124.
A192. Horne, B.D., et al., Early inpatient calculation of laboratory-based 30-day readmission risk scores empowers clinical risk modification during index hospitalization. Am Heart J, 2017. 185: p. 101-109.
A193. Itaya, T., et al., Assessment model to identify patients with stroke with a high possibility of discharge to home a retrospective cohort study. Stroke, 2017. 48(10): p. 2812-2818.
A194. Jamei, M., et al., Predicting all-cause risk of 30-day hospital readmission using artificial neural networks. PLoS ONE, 2017. 12(7).
A195. Jin, Y., T. Jin, and S.M. Lee, Automated Pressure Injury Risk Assessment System Incorporated Into an Electronic Health Record System. Nurs Res, 2017. 66(6): p. 462-472.
A196. Kaewprag, P., et al., Predictive models for pressure ulcers from intensive care unit electronic health records using Bayesian networks. BMC Med Inform Decis Mak, 2017. 17: p. 65.
A197. Karter, A.J., et al., Development and validation of a tool to identify patients with type 2 diabetes at high risk of hypoglycemia-related emergency department or hospital use. JAMA Intern Med, 2017. 177(10): p. 1461-1470.
A198. Kartoun, U., et al., The MELD-Plus: A generalizable prediction risk score in cirrhosis. PLoS ONE, 2017. 12(10).
A199. Kasbekar, P.U., P. Goel, and S.P. Jadhav, A decision tree analysis of diabetic foot amputation risk in Indian patients. Front Endocrinol, 2017. 8(FEB).
A200. Kessler, R.C., et al., Developing a practical suicide risk prediction model for targeting high-risk patients in the Veterans health Administration. Int J Methods Psychiatr Res, 2017. 26(3).
A201. Kim, E.H., et al., Prediction model for non-curative resection of endoscopic submucosal dissection in patients with early gastric cancer. Gastrointest Endosc, 2017. 85(5): p. 976-983.
A202. Li, H.Y., et al., A risk prediction score model for predicting occurrence of post-PCI vasovagal reflex syndrome: A single center study in Chinese population. J Geriatr Cardiol, 2017. 14(8): p. 509-514.
A203. Lin, K.J., et al., Prediction score for anticoagulation control quality among older adults. J Am Heart Assoc, 2017. 6(10).
A204. Lodhi, M.K., et al., Predicting Hospital Re-admissions from Nursing Care Data of Hospitalized Patients. Adv. data min., 2017. 2017: p. 181-193.
A205. Low, L.L., et al., FAM-FACE-SG: a score for risk stratification of frequent hospital admitters. BMC Med Inform Decis Mak, 2017. 17(1): p. 35.
A206. Lucas, J.E., et al., An electronic health record based model predicts statin adherence, LDL cholesterol, and cardiovascular disease in the United States Military Health System. PLoS ONE, 2017. 12(11).
A207. Makam, A.N., et al., Predicting 30-day pneumonia readmissions using electronic health record data. J Hosp Med, 2017. 12(4): p. 209-216.
A208. McKown, A.C., et al., Predicting Major Adverse Kidney Events among Critically Ill Adults Using the Electronic Health Record. J Med Syst, 2017. 41(10).
A209. Nakagawa, T., et al., Nomogram for predicting survival of postcystectomy recurrent urothelial carcinoma of the bladder. Urol Oncol Semin Orig Invest, 2017. 35(7): p. 457.e15-457.e21.
A210. Oliva, E.M., et al., Development and applications of the Veterans Health Administration's Stratification Tool for Opioid Risk Mitigation (STORM) to improve opioid safety and prevent overdose and suicide. Psychol Serv, 2017. 14(1): p. 34-49.
A211. Pan, L., et al., Machine learning applications for prediction of relapse in childhood acute lymphoblastic leukemia. Sci Rep, 2017. 7(1): p. 7402.
A212. Poppe, K.K., et al., Developing and validating a cardiovascular risk score for patients in the community with prior cardiovascular disease. Heart, 2017. 103(12): p. 917-922.
A213. Qiu, H., et al., Electronic Health Record Driven Prediction for Gestational Diabetes Mellitus in Early Pregnancy. Sci Rep, 2017. 7(1): p. 16417.
A214. Raman, J.D., et al., Preoperative nomogram to predict the likelihood of complications after radical nephroureterectomy. BJU Int, 2017. 119(2): p. 268-275.
A215. Rubin, D.J., et al., Predicting readmission risk of patients with diabetes hospitalized for cardiovascular disease: a retrospective cohort study. J Diabetes Complications, 2017. 31(8): p. 1332-1339.
A216. Sakhnini, A., et al., The derivation and validation of a simple model for predicting in-hospital mortality of acutely admitted patients to internal medicine wards. Medicine, 2017. 96(25).
A217. Schroeder, E.B., et al., Predicting the 6-month risk of severe hypoglycemia among adults with diabetes: Development and external validation of a prediction model. J Diabetes Complications, 2017. 31(7): p. 1158-1163.
A218. Tabak, Y.P., et al., Predicting Readmission at Early Hospitalization Using Electronic Clinical Data: An Early Readmission Risk Score. Med Care, 2017. 55(3): p. 267-275.
A219. Tuck, M.G., et al., A Comprehensive Index for Predicting Risk of Anemia from Patients' Diagnoses. Big Data, 2017. 5(1): p. 42-52.
A220. Viangteeravat, T., O. Akbilgic, and R.L. Davis, Analyzing Electronic Medical Records to Predict Risk of DIT (Death, Intubation, or Transfer to ICU) in Pediatric Respiratory Failure or Related Conditions. AMIA Summits Transl Sci Proc, 2017. 2017: p. 287-294.
A221. Weng, S.F., et al., Can machine-learning improve cardiovascular risk prediction using routine clinical data? PLoS ONE, 2017. 12(4): p. e0174944.
A222. Williams, B.A., et al., Clinical prediction model for time in therapeutic range while on warfarin in newly diagnosed atrial fibrillation. J Am Heart Assoc, 2017. 6(10).
A223. Zhang, Z. and Y. Hong, Development of a novel score for the prediction of hospital mortality in patients with severe sepsis: The use of electronic healthcare records with LASSO regression. Oncotarget, 2017. 8(30): p. 49637-49645.

## 2018

A224. Adelson, K., et al., Development of imminent mortality predictor for advanced cancer (IMPAC), a tool to predict short-term mortality in hospitalized patients with advanced cancer. J Oncol Pract, 2018. 14(3): p. e168-e175.
A225. Alderden, J., et al., Predicting Pressure Injury in Critical Care Patients: A Machine-Learning Model. Am J Crit Care, 2018. 27(6): p. 461-468.
A226. Arruda-Olson, A.M., et al., Leveraging the electronic health record to create an automated real-time prognostic tool for peripheral arterial disease. J Am Heart Assoc, 2018. 7(23).
A227. Avati, A., et al., Improving palliative care with deep learning. BMC Med Inform Decis Mak, 2018. 18: p. 122.
A228. Bertsimas, D., et al., Applied informatics decision support tool for mortality predictions in patients with cancer. JCO Clin Cancer Inform, 2018. 2018(2): p. 1-11.
A229. Bowen, G.S., et al., A Multivariable Prediction Model for Mortality in Individuals Admitted for Heart Failure. J Am Geriatr Soc, 2018. 66(5): p. 902-908.
A230. Brannon, E., et al., Towards a Learning Health System to Reduce Emergency Department Visits at a Population Level. AMIA Annu Symp Proc, 2018. 2018: p. 295-304.
A231. Brittan, M.S., et al., An Electronic Health Record Tool Designed to Improve Pediatric Hospital Discharge has Low Predictive Utility for Readmissions. J Hosp Med, 2018. 13(11): p. 779-782.
A232. Buchlak, Q.D., et al., Risk stratification in deep brain stimulation surgery: Development of an algorithm to predict patient discharge disposition with 91.9% accuracy. J Clin Neurosci, 2018. 57: p. 26-32.
A233. Burton, C.L., et al., Predicting surgical intervention in patients presenting with carpal tunnel syndrome in primary care. Clin Epidemiol, 2018. 10: p. 739-748.
A234. Calcaterra, S.L., et al., Prediction of Future Chronic Opioid Use Among Hospitalized Patients. J Gen Intern Med, 2018. 33(6): p. 898-905.
A235. Cappellari, M., et al., A nomogram to predict the probability of mortality after first-ever acute manifestations of cerebral small vessel disease. J Neurol Sci, 2018. 385: p. 92-95.
A236. Chan, D.X.H., et al., Development of the Combined Assessment of Risk Encountered in Surgery (CARES) surgical risk calculator for prediction of postsurgical mortality and need for intensive care unit admission risk: A single-center retrospective study. BMJ Open, 2018. 8(3).
A237. Chen, D., et al., Postoperative bleeding risk prediction for patients undergoing colorectal surgery. Surgery, 2018. 164(6): p. 1209-1216.
A238. Chen, L., et al., Performance and validation of a simplified postoperative atrial fibrillation risk score. PACE Pacing Clin Electrophysiol, 2018. 41(9): p. 1136-1142.
A239. Cho, U., et al., Prognostic value of systemic inflammatory markers and development of a nomogram in breast cancer. PLoS ONE, 2018. 13(7).
A240. Choi, S.B., et al., Ten-year prediction of suicide death using Cox regression and machine learning in a nationwide retrospective cohort study in South Korea. J Affective Disord, 2018. 231: p. 8-14.
A241. Choi, Y., et al., A dynamic risk model for inpatient falls. Am J Health-Syst Pharm, 2018. 75(17): p. 1293-1303.
A242. Corradi, J.P., et al., Prediction of Incident Delirium Using a Random Forest classifier. J Med Syst, 2018. 42(12).
A243. Dagliati, A., et al., Machine Learning Methods to Predict Diabetes Complications. J Diabetes Sci Technol, 2018. 12(2): p. 295-302.
A244. Delahanty, R.J., D. Kaufman, and S.S. Jones, Development and Evaluation of an Automated Machine Learning Algorithm for In-Hospital Mortality Risk Adjustment Among Critical Care Patients. Crit Care Med, 2018. 46(6): p. e481-e488.
A245. Ding, X., et al., Designing risk prediction models for ambulatory no-shows across different specialties and clinics. J Am Med Informatics Assoc, 2018. 25(8): p. 924-930.
A246. Dreijer, A.R., et al., Development of a clinical prediction model for an international normalised ratio ≥ 4·5 in hospitalised patients using vitamin K antagonists. Br J Haematol, 2018. 181(1): p. 102-110.
A247. Dziadzko, M.A., et al., Multicenter derivation and validation of an early warning score for acute respiratory failure or death in the hospital. Crit Care, 2018. 22(1).
A248. Elfiky, A.A., et al., Development and Application of a Machine Learning Approach to Assess Short-term Mortality Risk Among Patients With Cancer Starting Chemotherapy. JAMA Netw Open, 2018. 1(3): p. e180926.
A249. Faisal, M., et al., A comparison of logistic regression models with alternative machine learning methods to predict the risk of in-hospital mortality in emergency medical admissions via external validation. Health Inform J, 2018: p. 1460458218813600.
A250. Faisal, M., et al., Development and external validation of an automated computer-aided risk score for predicting sepsis in emergency medical admissions using the patient's first electronically recorded vital signs and blood test results. Crit Care Med, 2018. 46(4): p. 612-618.
A251. Faisal, M., et al., Development and validation of a novel computer-aided score to predict the risk of in-hospital mortality for acutely ill medical admissions in two acute hospitals using their first electronically recorded blood test results and vital signs: A cross-sectional study. BMJ Open, 2018. 8(12).
A252. Franklin, J.M., et al., Time to Filling of New Prescriptions for Chronic Disease Medications Among a Cohort of Elderly Patients in the USA. J Gen Intern Med, 2018. 33(11): p. 1877-1884.
A253. Gilbert, T., et al., Development and validation of a Hospital Frailty Risk Score focusing on older people in acute care settings using electronic hospital records: an observational study. Lancet, 2018. 391(10132): p. 1775-1782.
A254. Glanz, J.M., et al., Prediction Model for Two-Year Risk of Opioid Overdose Among Patients Prescribed Chronic Opioid Therapy. J Gen Intern Med, 2018: p. 1-8.
A255. Grinspan, Z.M., et al., Predicting frequent emergency department use among children with epilepsy: A retrospective cohort study using electronic health data from 2 centers. Epilepsia, 2018. 59(1): p. 155-169.
A256. Guo, Y., et al., Assessing Statewide All-Cause Future One-Year Mortality: Prospective Study With Implications for Quality of Life, Resource Utilization, and Medical Futility. J Med Internet Res, 2018. 20(6): p. e10311.
A257. Hatipoğlu, U., et al., Predicting 30-Day All-Cause Readmission Risk for Subjects Admitted With Pneumonia at the Point of Care. Respir Care, 2018. 63(1): p. 43-49.
A258. Hong, J.C., et al., Predicting emergency visits and hospital admissions during radiation and chemoradiation: An internally validated pretreatment machine learning algorithm. JCO Clin Cancer Inform, 2018. 2018(2): p. 1-11.
A259. Horta, A.B., et al., Clinical decision support tool for Co-management signalling. Int J Med Informatics, 2018. 113: p. 56-62.
A260. Huang, Z., et al., Analysis of a large data set to identify predictors of blood transfusion in primary total hip and knee arthroplasty. Transfusion, 2018. 58(8): p. 1855-1862.
A261. Hur, E.Y., et al., Development and evaluation of the automated risk assessment system for multidrug-resistant organisms (autoRAS-MDRO). J Hosp Infect, 2018. 98(2): p. 202-211.
A262. Kan, H.J., et al., Factors associated with physicians' prescriptions for rheumatoid arthritis drugs not filled by patients. Arthritis Res Ther, 2018. 20(1).
A263. Khojandi, A., et al., Prediction of Sepsis and In-Hospital Mortality Using Electronic Health Records. Methods Inf Med, 2018. 57(4): p. 185-193.
A264. Kleiman, R.S., et al., Using Machine Learning Algorithms to Predict Risk for Development of Calciphylaxis in Patients with Chronic Kidney Disease. AMIA Summits Transl Sci Proc, 2018. 2017: p. 139-146.
A265. Koyner, J.L., et al., The development of a machine learning inpatient acute kidney injury prediction model. Crit Care Med, 2018. 46(7): p. 1070-1077.
A266. Kraaijvanger, N., et al., Development and validation of an admission prediction tool for emergency departments in the Netherlands. Emerg Med J, 2018. 35(8): p. 464-470.
A267. Lagu, T., et al., Derivation and validation of an in-hospital mortality prediction model suitable for profiling hospital performance in heart failure. J Am Heart Assoc, 2018. 7(4).
A268. Lee, J.Y., H.A. Park, and E. Chung, Use of electronic critical care flow sheet data to predict unplanned extubation in ICUs. Int J Med Informatics, 2018. 117: p. 6-12.
A269. Lee, M.J., et al., Can we predict when to start renal replacement therapy in patients with chronic kidney disease using 6 months of clinical data? PLoS ONE, 2018. 13(10).
A270. Levine, M., et al., Hypoglycemia and lactic acidosis outperform King’s College criteria for predicting death or transplant in acetaminophen toxic patients. Clin Toxicol, 2018. 56(7): p. 622-625.
A271. Li, T., et al., Predicting Neonatal Encephalopathy From Maternal Data in Electronic Medical Records. AMIA Summits Transl Sci Proc, 2018. 2017: p. 359-368.
A272. Lim, Y.J., et al., A novel prognostic nomogram for predicting risks of distant failure in patients with invasive breast cancer following postoperative adjuvant radiotherapy. Cancer Res Treat, 2018. 50(4): p. 1140-1148.
A273. Lin, H., et al., Prediction of myopia development among Chinese school-aged children using refraction data from electronic medical records: A retrospective, multicentre machine learning study. PLoS Med, 2018. 15(11).
A274. Liu, L., et al., An interpretable boosting model to predict side effects of analgesics for osteoarthritis. BMC Syst Biol, 2018. 12: p. 105.
A275. Martínez-Laguna, D., et al., Fracture risk in type 2 diabetic patients: A clinical prediction tool based on a large population-based cohort. PLoS ONE, 2018. 13(9).
A276. McAuliffe, L., et al., Development and validation of a transitions-of-care pharmacist tool to predict potentially avoidable 30-day readmissions. Am J Health-Syst Pharm, 2018. 75(3): p. 111-119.
A277. McNairy, M.L., et al., Predicting death and lost to follow-up among adults initiating antiretroviral therapy in resource-limited settings: Derivation and external validation of a risk score in Haiti. Plos One, 2018. 13(8).
A278. Mohamadlou, H., et al., Prediction of Acute Kidney Injury With a Machine Learning Algorithm Using Electronic Health Record Data. Can J Kidney Health Dis, 2018. 5.
A279. Mohammadi, I., et al., Data Analytics and Modeling for Appointment No-show in Community Health Centers. J Prim Care Community Health, 2018. 9: p. 2150132718811692.
A280. Moon, K.J., et al., Development and validation of an automated delirium risk assessment system (Auto-DelRAS) implemented in the electronic health record system. Int J Nurs Stud, 2018. 77: p. 46-53.
A281. Nguyen, O.K., et al., Predicting 30-Day Hospital Readmissions in Acute Myocardial Infarction: The AMI "READMITS" (Renal Function, Elevated Brain Natriuretic Peptide, Age, Diabetes Mellitus, Nonmale Sex, Intervention with Timely Percutaneous Coronary Intervention, and Low Systolic Blood Pressure) Score. J Am Heart Assoc, 2018. 7(8).
A282. Oh, J., et al., A Generalizable, Data-Driven Approach to Predict Daily Risk of Clostridium difficile Infection at Two Large Academic Health Centers. Infect Control Hosp Epidemiol, 2018. 39(4): p. 425-433.
A283. Ortega-Loubon, C., et al., Predictors of Postoperative Acute Kidney Injury after Coronary Artery Bypass Graft Surgery. Braz J Cardiovasc Surg, 2018. 33(4): p. 323-329.
A284. Pakbin, A., et al., Prediction of ICU Readmissions Using Data at Patient Discharge. Conf Proc IEEE Eng Med Biol Soc, 2018. 2018: p. 4932-4935.
A285. Perveen, S., et al., A Systematic Machine Learning Based Approach for the Diagnosis of Non-Alcoholic Fatty Liver Disease Risk and Progression. Sci Rep, 2018. 8(1): p. 2112.
A286. Pimentel, A., et al., Screening diabetes mellitus 2 based on electronic health records using temporal features. Health Informatics J, 2018. 24(2): p. 194-205.
A287. Rahimian, F., et al., Predicting the risk of emergency admission with machine learning: Development and validation using linked electronic health records. PLoS Med, 2018. 15(11).
A288. Reber, K.C., et al., Development of a risk assessment tool for osteoporotic fracture prevention: A claims data approach. Bone, 2018. 110: p. 170-176.
A289. Reddy, B.K. and D. Delen, Predicting hospital readmission for lupus patients: An RNN-LSTM-based deep-learning methodology. Comput Biol Med, 2018. 101: p. 199-209.
A290. Rojas, J.C., et al., Predicting intensive care unit readmission with machine learning using electronic health record data. Ann Am Thorac Soc, 2018. 15(7): p. 846-853.
A291. Rubin, J., et al., An ensemble boosting model for predicting transfer to the pediatric intensive care unit. Int J Med Informatics, 2018. 112: p. 15-20.
A292. Rubin, K.H., et al., A New Fracture Risk Assessment Tool (FREM) Based on Public Health Registries. J Bone Miner Res, 2018. 33(11): p. 1967-1979.
A293. Sahni, N., G. Simon, and R. Arora, Development and Validation of Machine Learning Models for Prediction of 1-Year Mortality Utilizing Electronic Medical Record Data Available at the End of Hospitalization in Multicondition Patients: a Proof-of-Concept Study. J Gen Intern Med, 2018. 33(6): p. 921-928.
A294. Saqib, M., Y. Sha, and M.D. Wang, Early Prediction of Sepsis in EMR Records Using Traditional ML Techniques and Deep Learning LSTM Networks. Conf Proc IEEE Eng Med Biol Soc, 2018. 2018: p. 4038-4041.
A295. Satchidanand, N., et al., Development of a Risk Tool to Support Discussions of Care for Older Adults Admitted to the ICU With Pneumonia. Am J Hosp Palliat Care, 2018. 35(9): p. 1201-1206.
A296. Schmidt, C.R., et al., Development and prospective validation of a model estimating risk of readmission in cancer patients. J Surg Oncol, 2018. 117(6): p. 1113-1118.
A297. Schwartz, N., A. Sakhnini, and N. Bisharat, Predictive modeling of inpatient mortality in departments of internal medicine. Intern Emerg Med, 2018. 13(2): p. 205-211.
A298. Siregar, N.N., et al., Seventy-two hour mortality prediction model in patients with diabetic ketoacidosis: A retrospective cohort study. J ASEAN Fed Endocr Soc, 2018. 33(2): p. 124-129.
A299. Stone, A.V., et al., Nutritional markers may identify patients with greater risk of re-admission after geriatric hip fractures. Int Orthop, 2018. 42(2): p. 231-238.
A300. Suchting, R., et al., A data science approach to predicting patient aggressive events in a psychiatric hospital. Psychiatry Res, 2018. 268: p. 217-222.
A301. Wang, H., et al., Predicting Hospital Readmission via Cost-Sensitive Deep Learning. IEEE/ACM Trans Comput Biol Bioinform, 2018. 15(6): p. 1968-1978.
A302. Winterstein, A.G., et al., Development and validation of an automated algorithm for identifying patients at high risk for drug-induced hypoglycemia. Am J Health-Syst Pharm, 2018. 75(21): p. 1714-1728.
A303. Wojtusiak, J., E. Elashkar, and R. Mogharab Nia, C-LACE2: computational risk assessment tool for 30-day post hospital discharge mortality. Health Technol, 2018. 8(5): p. 341-351.
A304. Wong, A., et al., Development and Validation of an Electronic Health Record-Based Machine Learning Model to Estimate Delirium Risk in Newly Hospitalized Patients Without Known Cognitive Impairment. JAMA Netw Open, 2018. 1(4): p. e181018.
A305. Wu, Y., et al., Quantifying predictive capability of electronic health records for the most harmful breast cancer. 2018.
A306. Ye, C., et al., Prediction of Incident Hypertension Within the Next Year: Prospective Study Using Statewide Electronic Health Records and Machine Learning. J Med Internet Res, 2018. 20(1): p. e22.
A307. Young, J.B., et al., Development of predictive risk models for major adverse cardiovascular events among patients with type 2 diabetes mellitus using health insurance claims data. Cardiovasc Diabetol, 2018. 17(1).

## 2019

A308. Akbilgic, O., et al., Machine Learning to Identify Dialysis Patients at High Death Risk. Kidney Intl Rep, 2019. 4(9): p. 1219-1229.
A309. Alavifard, S., et al., Derivation and validation of a model predicting the likelihood of vaginal birth following labour induction. BMC Pregnancy Childbirth, 2019. 19(1).
A310. Ashfaq, A., et al., Readmission prediction using deep learning on electronic health records. J Biomed Informatics, 2019. 97.
A311. Atlantis, E., et al., A predictive model for non-completion of an intensive specialist obesity service in a public hospital: a case-control study. BMC Health Serv Res, 2019. 19(1): p. 748.
A312. Avtaar Singh, S.S., et al., PREDICTA: A Model to Predict Primary Graft Dysfunction After Adult Heart Transplantation in the United Kingdom. J Card Fail, 2019.
A313. Baxter, S.L., et al., Machine Learning-Based Predictive Modeling of Surgical Intervention in Glaucoma Using Systemic Data From Electronic Health Records. Am J Ophthalmol, 2019. 208: p. 30-40.
A314. Beeksma, M., et al., Predicting life expectancy with a long short-term memory recurrent neural network using electronic medical records. BMC Med Inform Decis Mak, 2019. 19(1): p. 36.
A315. Blanc, A.L., et al., Development of a predictive score for potentially avoidable hospital readmissions for general internal medicine patients. PLoS ONE, 2019. 14(7).
A316. Blom, M.C., et al., Training machine learning models to predict 30-day mortality in patients discharged from the emergency department: A retrospective, population-based registry study. BMJ Open, 2019. 9(8).
A317. Bloom, C.I., et al., Predicting COPD 1-year mortality using prognostic predictors routinely measured in primary care. BMC Med, 2019. 17(1).
A318. Carmen, R., et al., The role of specialized hospital units in infection and mortality risk reduction among patients with hematological cancers. PLoS ONE, 2019. 14(3).
A319. Chang, H.Y., et al., A predictive risk model for nonfatal opioid overdose in a statewide population of buprenorphine patients. Drug Alcohol Depend, 2019. 201: p. 127-133.
A320. Chaudhary, M.A., et al., Development and Validation of a Bedside Risk Assessment for Sustained Prescription Opioid Use after Surgery. JAMA Netw Open, 2019. 2(7).
A321. Chiew, C.J., et al., Utilizing Machine Learning Methods for Preoperative Prediction of Postsurgical Mortality and Intensive Care Unit Admission. Ann Surg, 2019.
A322. Cho, I., et al., Novel Approach to Inpatient Fall Risk Prediction and Its Cross-Site Validation Using Time-Variant Data. J Med Internet Res, 2019. 21(2): p. e11505.
A323. Choi, B.G., et al., Machine learning for the prediction of new-onset diabetes mellitus during 5-year follow-up in non-diabetic patients with cardiovascular risks. Yonsei Med J, 2019. 60(2): p. 191-199.
A324. Clarke, C.L., et al., Predictors of long-term survival among high-grade serous ovarian cancer patients. Cancer Epidemiol Biomarkers Prev, 2019. 28(5): p. 996-999.
A325. Cramer, E.M., et al., Predicting the Incidence of Pressure Ulcers in the Intensive Care Unit Using Machine Learning. EGEMS (Wash DC), 2019. 7(1): p. 49.
A326. Daghistani, T.A., et al., Predictors of in-hospital length of stay among cardiac patients: A machine learning approach. Int J Cardiol, 2019. 288: p. 140-147.
A327. Danilov, G., et al., Prediction of Postoperative Hospital Stay with Deep Learning Based on 101 654 Operative Reports in Neurosurgery. Stud Health Technol Inform, 2019. 258: p. 125-129.
A328. Edgcomb, J., et al., High-Risk Phenotypes of Early Psychiatric Readmission in Bipolar Disorder With Comorbid Medical Illness. Psychosomatics, 2019.
A329. Fouks, Y., et al., Surgical Intervention in Patients with Tubo-Ovarian Abscess: Clinical Predictors and a Simple Risk Score. J Minimally Invasive Gynecol, 2019. 26(3): p. 535-543.
A330. Franckowiak, T.M., J.N. Raub, and R. Yost, Derivation and validation of a hospital all-cause 30-day readmission index. Am J Health-Syst Pharm, 2019. 76(7): p. 436-443.
A331. Galaznik, A., et al., Predicting Outcomes in Patients With Diffuse Large B-Cell Lymphoma Treated With Standard of Care. Cancer Informatics, 2019. 18.
A332. Gao, C., et al., Deep learning predicts extreme preterm birth from electronic health records. J Biomed Informatics, 2019. 100.
A333. Gensheimer, M.F., et al., Automated Survival Prediction in Metastatic Cancer Patients Using High-Dimensional Electronic Medical Record Data. J Natl Cancer Inst, 2019. 111(6): p. 568-574.
A334. Giannini, H.M., et al., A Machine Learning Algorithm to Predict Severe Sepsis and Septic Shock: Development, Implementation, and Impact on Clinical Practice. Crit Care Med, 2019. 47(11): p. 1485-1492.
A335. Ginanjar, E., et al., Predictors of 30-day Mortality in ST-Elevation Myocardial Infarction (STEMI) Patients. Acta Med Indones, 2019. 51(3): p. 238-244.
A336. Goltz, D.E., et al., A Novel Risk Calculator Predicts 90-Day Readmission Following Total Joint Arthroplasty. J Bone Jt Surg Am Vol, 2019. 101(6): p. 547-556.
A337. Goltz, D.E., et al., A Weighted Index of Elixhauser Comorbidities for Predicting 90-day Readmission After Total Joint Arthroplasty. J Arthroplasty, 2019. 34(5): p. 857-864.
A338. Gupta, A., T. Liu, and S. Shepherd, Clinical decision support system to assess the risk of sepsis using Tree Augmented Bayesian networks and electronic medical record data. Health Inform J, 2019: p. 1460458219852872.
A339. Hall, R.K., et al., A Novel Approach to Developing a Discordance Index for Older Adults with Chronic Kidney Disease. J Gerontol A Biol Sci Med Sci, 2019.
A340. Hammond, R., et al., Predicting childhood obesity using electronic health records and publicly available data. PLoS ONE, 2019. 14(4).
A341. Hester, G.Z., et al., Identifying patients with Kawasaki disease safe for early discharge: Development of a risk prediction model at a US children’s hospital. Hosp Pediatr, 2019. 9(10): p. 749-756.
A342. Hill, B.L., et al., An automated machine learning-based model predicts postoperative mortality using readily-extractable preoperative electronic health record data. Br J Anaesth, 2019. 123(6): p. 877-886.
A343. Hincapie-Castillo, J.M., et al., Development of a predictive model for drug-associated QT prolongation in the inpatient setting using electronic health record data. Am J Health-Syst Pharm, 2019. 76(14): p. 1059-1070.
A344. Hung, C.Y., et al., Development of an intelligent decision support system for ischemic stroke risk assessment in a population-based electronic health record database. PLoS ONE, 2019. 14(3).
A345. Hur, E.Y., et al., Development and Evaluation of the Automated Risk Assessment System for Catheter-Associated Urinary Tract Infection. Comput Inform Nurs, 2019. 37(9): p. 463-472.
A346. Hyun, S., et al., Prediction Model for Hospital-Acquired Pressure Ulcer Development: Retrospective Cohort Study. JMIR Med Inform, 2019. 7(3): p. e13785.
A347. Jang, D.H., et al., Developing neural network models for early detection of cardiac arrest in emergency department. Am J Emerg Med, 2019.
A348. Jauk, S., et al., Development of a Machine Learning Model Predicting an ICU Admission for Patients with Elective Surgery and Its Prospective Validation in Clinical Practice. Stud Health Technol Inform, 2019. 264: p. 173-177.
A349. Jeon, N., et al., Development and validation of an automated algorithm for identifying patients at higher risk for drug-induced acute kidney injury. Am J Health-Syst Pharm, 2019. 76(10): p. 654-666.
A350. Jhee, J.H., et al., Prediction model development of late-onset preeclampsia using machine learning-based methods. PLoS ONE, 2019. 14(8).
A351. Jiang, X., et al., Prognostic nomogram for acute pancreatitis patients: An analysis of publicly electronic healthcare records in intensive care unit. J Crit Care, 2019. 50: p. 213-220.
A352. Jung, H., H.A. Park, and H. Hwang, Improving Prediction of Fall Risk Using Electronic Health Record Data With Various Types and Sources at Multiple Times. Comput Inform Nurs, 2019.
A353. Kaji, D.A., et al., An attention based deep learning model of clinical events in the intensive care unit. PLoS ONE, 2019. 14(2).
A354. Karhade, A.V., et al., Machine learning for prediction of sustained opioid prescription after anterior cervical discectomy and fusion. Spine J, 2019. 19(6): p. 976-983.
A355. Karhade, A.V., J.H. Schwab, and H.S. Bedair, Development of Machine Learning Algorithms for Prediction of Sustained Postoperative Opioid Prescriptions After Total Hip Arthroplasty. J Arthroplasty, 2019. 34(10): p. 2272-2277.e1.
A356. Koning, N.R., et al., Identification of children at risk for mental health problems in primary care—Development of a prediction model with routine health care data. EClinicalMedicine, 2019. 15: p. 89-97.
A357. Krakower, D.S., et al., Development and validation of an automated HIV prediction algorithm to identify candidates for pre-exposure prophylaxis: a modelling study. Lancet HIV, 2019. 6(10): p. e696-e704.
A358. Le, S., et al., Pediatric Severe Sepsis Prediction Using Machine Learning. Front Pediatr, 2019. 7.
A359. Leary, J.C., et al., Developing prediction models for 30-day unplanned readmission among children with medical complexity. Hosp Pediatr, 2019. 9(3): p. 201-208.
A360. Lee, J.A., et al., Algorithm to Predict the Outcome of Microvascular Decompression for Hemifacial Spasm: A Data-Mining Analysis Using a Decision Tree. World Neurosurg, 2019. 125: p. e797-e806.
A361. Lee, S.Y., et al., Prediction of emergency department patient disposition decision for proactive resource allocation for admission. 2019.
A362. Li, B.Y., et al., Using Machine learning and the electronic health record to predict complicated clostridium difficile infection. Open Forum Infect Dis, 2019. 6(5).
A363. Li, H., et al., Decision tree model for predicting in-hospital cardiac arrest among patients admitted with acute coronary syndrome. Clin Cardiol, 2019.
A364. Li, X.J., et al., Surgeon-Specific Risk Stratification Model for Early Complications After Complex Adult Spinal Deformity Surgery. Spine Deform, 2019.
A365. Li, Y., et al., Development and validation of a dynamic inpatient risk prediction model for clinically significant hypokalemia using electronic health record data. Am J Health-Syst Pharm, 2019. 76(5): p. 301-311.
A366. Liu, L., et al., Mining patient-specific and contextual data with machine learning technologies to predict cancellation of children's surgery. Int J Med Informatics, 2019. 129: p. 234-241.
A367. Low, S., et al., Electronic health records accurately predict renal replacement therapy in acute kidney injury. BMC Nephrol, 2019. 20(1).
A368. Lu, H.X., et al., Development and validation of a novel predictive score for sepsis risk among trauma patients. World J Emerg Surg, 2019. 14: p. 11.
A369. Makino, M., et al., Artificial intelligence predicts the progression of diabetic kidney disease using big data machine learning. Sci. rep., 2019. 9(1): p. 11862.
A370. Maltenfort, M.G., Y. Chen, and C.B. Forrest, Prediction of 30-day pediatric unplanned hospitalizations using the Johns Hopkins Adjusted Clinical Groups risk adjustment system. PLoS ONE, 2019. 14(8).
A371. Marcus, J.L., et al., Use of electronic health record data and machine learning to identify candidates for HIV pre-exposure prophylaxis: a modelling study. Lancet HIV, 2019. 6(10): p. e688-e695.
A372. Martin, A., et al., Development and validation of an asthma exacerbation prediction model using electronic health record (EHR) data. J Asthma, 2019.
A373. Menger, V., et al., Machine Learning Approach to Inpatient Violence Risk Assessment Using Routinely Collected Clinical Notes in Electronic Health Records. JAMA Netw Open, 2019. 2(7).
A374. Mo, X., et al., Early and accurate prediction of clinical response to methotrexate treatment in juvenile idiopathic arthritis using machine learning. Front Pharmacol, 2019. 10.
A375. Mohammadi, R., et al., Learning to Identify Patients at Risk of Uncontrolled Hypertension Using Electronic Health Records Data. AMIA Summits Transl Sci Proc, 2019. 2019: p. 533-542.
A376. Morsy, A.M., et al., Age and Tumor Location Predict Survival in Nonmetastatic Osteosarcoma in Upper Egypt. J Pediatr Hematol Oncol, 2019.
A377. Muñoz, M.A., et al., Predicting medication-associated altered mental status in hospitalized patients: Development and validation of a risk model. Am J Health-Syst Pharm, 2019. 76(13): p. 953-963.
A378. Noe, M.H., et al., Development and Validation of a Risk Prediction Model for In-Hospital Mortality among Patients with Stevens-Johnson Syndrome/Toxic Epidermal Necrolysis - ABCD-10. JAMA Dermatol, 2019. 155(4): p. 448-454.
A379. Norgeot, B., et al., Assessment of a Deep Learning Model Based on Electronic Health Record Data to Forecast Clinical Outcomes in Patients With Rheumatoid Arthritis. JAMA Netw Open, 2019. 2(3): p. e190606.
A380. Oseran, A.S., et al., A “Hospital-Day-1” Model to Predict the Risk of Discharge to a Skilled Nursing Facility. J Am Med Dir Assoc, 2019. 20(6): p. 689-695.e5.
A381. Oshiro, C.E.S., et al., Fall Ascertainment and Development of a Risk Prediction Model Using Electronic Medical Records. J Am Geriatr Soc, 2019.
A382. Paik, E.S., et al., Prediction of survival outcomes in patients with epithelial ovarian cancer using machine learning methods. J Gynecol Oncol, 2019. 30(4).
A383. Pan, H., et al., Risk factors associated with prolonged air leak after video-assisted thoracic surgery pulmonary resection: A predictive model and meta-analysis. Ann Transl Med, 2019. 7(5).
A384. Parikh, R.B., et al., Machine Learning Approaches to Predict 6-Month Mortality among Patients with Cancer. JAMA Netw Open, 2019.
A385. Parker, C.A., et al., Predicting hospital admission at the emergency department triage: A novel prediction model. Am J Emerg Med, 2019. 37(8): p. 1498-1504.
A386. Parthipan, A., et al., Predicting inadequate postoperative pain management in depressed patients: A machine learning approach. PLoS ONE, 2019. 14(2).
A387. Patterson, B.W., et al., Training and Interpreting Machine Learning Algorithms to Evaluate Fall Risk After Emergency Department Visits. Med Care, 2019. 57(7): p. 560-566.
A388. Pauly, V., et al., Predictive risk score for unplanned 30-day rehospitalizations in the French universal health care system based on a medico-administrative database. PLoS ONE, 2019. 14(3).
A389. Pearce, C., et al., POLAR Diversion: Using General Practice Data to Calculate Risk of Emergency Department Presentation at the Time of Consultation. Applied Clinical Informatics, 2019. 10(1): p. 151-157.
A390. Perveen, S., et al., Prognostic Modeling and Prevention of Diabetes Using Machine Learning Technique. Sci Rep, 2019. 9(1): p. 13805.
A391. Pieszko, K., et al., Predicting long-term mortality after acute coronary syndrome using machine learning techniques and hematological markers. Dis Markers, 2019. 2019.
A392. Quan, J., et al., Risk Prediction Scores for Mortality, Cerebrovascular, and Heart Disease among Chinese People with Type 2 Diabetes. J Clin Endocrinol Metab, 2019. 104(12): p. 5823-5830.
A393. Reddy, B.K., D. Delen, and R.K. Agrawal, Predicting and explaining inflammation in Crohn's disease patients using predictive analytics methods and electronic medical record data. Health Informatics J, 2019. 25(4): p. 1201-1218.
A394. Restrepo-Escobar, M., P.A. Granda-Carvajal, and F. Jaimes, Development and Internal Validation of a Prediction Model to Estimate the Probability of Needing Aggressive Immunosuppressive Therapy With Cytostatics in de Novo Lupus Nephritis Patients. Reumatol Clin, 2019. 15(1): p. 27-33.
A395. Ross, E.G., et al., Predicting Future Cardiovascular Events in Patients With Peripheral Artery Disease Using Electronic Health Record Data. Circ Cardiovasc Qual Outcomes, 2019. 12(3): p. e004741.
A396. Samuels-Kalow, M.E., et al., A Predictive Model for Identification of Children at Risk of Subsequent High-Frequency Utilization of the Emergency Department for Asthma. Pediatr Emerg Care, 2019.
A397. Sanaee, M.S., et al., Urinary tract infection after clean-contaminated pelvic surgery: a retrospective cohort study and prediction model. Int Urogynecol J, 2019.
A398. Simonov, M., et al., A simple real-time model for predicting acute kidney injury in hospitalized patients in the US: A descriptive modeling study. PLoS Med, 2019. 16(7).
A399. Slieker, F.J.B., R. de Bree, and E.M. Van Cann, Predicting individualized mortality probabilities for patients with squamous cell carcinoma of the maxilla: Novel models with clinical and histopathological predictors. Head Neck, 2019. 41(10): p. 3584-3593.
A400. Soong, J.T.Y., et al., Dr Foster global frailty score: An international retrospective observational study developing and validating a risk prediction model for hospitalised older persons from administrative data sets. BMJ Open, 2019. 9(6).
A401. Tan, B.Y., et al., Electronic medical record-based model to predict the risk of 90-day readmission for patients with heart failure. BMC Med Inform Decis Mak, 2019. 19(1): p. 193.
A402. Tng, A.R.K., et al., Validation of the failure to maturation equation and proposal for a novel scoring system for arteriovenous fistula maturation in multiethnic Asian haemodialysis patients. J Vasc Access, 2019.
A403. Tsur, A., et al., Development and validation of a machine learning model for prediction of shoulder dystocia. Ultrasound Obstet Gynecol, 2019.
A404. Wang, H.H., et al., Assessment of Deep Learning Using Nonimaging Information and Sequential Medical Records to Develop a Prediction Model for Nonmelanoma Skin Cancer. JAMA Dermatol, 2019.
A405. Wang, L., et al., Development and Validation of a Deep Learning Algorithm for Mortality Prediction in Selecting Patients with Dementia for Earlier Palliative Care Interventions. JAMA Netw Open, 2019.
A406. Wang, S., J. Pathak, and Y. Zhang, Using Electronic Health Records and Machine Learning to Predict Postpartum Depression. Stud Health Technol Inform, 2019. 264: p. 888-892.
A407. Wang, X., et al., Prediction of the 1-Year Risk of Incident Lung Cancer: Prospective Study Using Electronic Health Records from the State of Maine. J Med Internet Res, 2019. 21(5): p. e13260.
A408. Wegier, P., et al., MHOMR: A feasibility study of an automated system for identifying inpatients having an elevated risk of 1-year mortality. BMJ Qual Saf, 2019.
A409. Wolf, A.E., et al., Pediatric acute myocarditis: Predicting hemodynamic compromise at presentation to health care. Hosp Pediatr, 2019. 9(6): p. 455-459.
A410. Wynn, J.L. and R.A. Polin, A neonatal sequential organ failure assessment score predicts mortality to late-onset sepsis in preterm very low birth weight infants. Pediatr Res, 2019.
A411. Wysham, C.H., et al., Development of risk models for major adverse chronic renal outcomes among patients with type 2 diabetes mellitus using insurance claims: a retrospective observational study. Curr Med Res Opin, 2019: p. 1.
A412. Xie, F., et al., Novel model for predicting inpatient mortality after emergency admission to hospital in Singapore: Retrospective observational study. BMJ Open, 2019. 9(9).
A413. Xu, F.B., et al., Derivation and validation of a prediction score for acute kidney injury secondary to acute myocardial infarction in Chinese patients. BMC Nephrol, 2019. 20(1).
A414. Yang, P.S., et al., A Novel Prediction Model for Bloodstream Infections in Hepatobiliary-Pancreatic Surgery Patients. World J Surg, 2019. 43(5): p. 1294-1302.
A415. Yee, C.R., et al., A Data-Driven Approach to Predicting Septic Shock in the Intensive Care Unit. Biomed. inform. insights, 2019. 11: p. 1178222619885147.
A416. You, Y., et al., Nomogram for predicting postoperative pancreatic fistula. HPB, 2019.
A417. Yu, D., et al., Development and validation of prediction models to estimate risk of primary total hip and knee replacements using data from the UK: Two prospective open cohorts using the UK Clinical Practice Research Datalink. Ann Rheum Dis, 2019. 78(1): p. 91-99.
A418. Zeiberg, D., et al., Machine learning for patient risk stratification for acute respiratory distress syndrome. PLoS ONE, 2019. 14(3).
A419. Zhang, M., et al., An interactive nomogram to predict healthcare-associated infections in ICU patients: A multicenter study in GuiZhou Province, China. PLoS ONE, 2019. 14(7).
A420. Zhang, S., et al., Predicting Recurrent Hypertensive Intracerebral Hemorrhage: Derivation and Validation of a Risk-Scoring Model Based on Clinical Characteristics. World Neurosurg, 2019. 127: p. e162-e171.
A421. Zhao, J., S. Gu, and A. McDermaid, Predicting outcomes of chronic kidney disease from EMR data based on Random Forest Regression. Math Biosci, 2019. 310: p. 24-30.
A422. Zhou, J., et al., A simple risk score for prediction of sepsis associated-acute kidney injury in critically ill patients. J Nephrol, 2019. 32(6): p. 947-956.
